# The impact of the COVID-19 pandemic on the mental health and wellbeing of UK healthcare workers

**DOI:** 10.1101/2020.10.23.20218396

**Authors:** James Gilleen, Aida Santaolalla, Lorena Valdearenas, Clara Salice, Montserrat Fusté

## Abstract

**Background:** There is an urgent need to understand the psychological impact the COVID-19 pandemic has had on UK healthcare workers (HCW).

**Aims:** To reveal risk and protective factors associated with poor mental wellbeing of HCW working during the COVID-19 pandemic in the UK.

**Method:** 2773 UK HCWs completed a survey between 22^nd^April and 10^th^ May 2020 containing scales measuring anxiety, depression, PTSD, and stress, and questions about roles and COVID-19-related factors including workplace preparation and risk management. Respondents were classified as high or low symptomatic on each mental health scale and logistic regression revealed risk and protective factors associated with each outcome. Change in wellbeing from pre to during COVID-19 was also quantified.

**Results:** A large proportion of UK HCW had high mental health symptoms. ‘*Fixed’* risk factors of poor mental health included being female, being ‘frontline’, pre-existing mental health diagnoses, and experience of stressful/traumatic events. An additional set of ‘*controllable’* factors also significantly increased risk: PPE availability, workload, lack of COVID-19 preparation and training, and insufficient communication of clinical procedures. Resilience and sharing stress reduced risk, as did ethical support for those making treatment decisions. Allied HCW and managers were at elevated risk of high symptoms particularly PTSD. Wellbeing, especially of frontline workers, had significantly worsened compared to before COVID-19.

**Conclusions:** Poor mental wellbeing was prevalent in HCW during the UK COVID-19 response. A number of controllable factors should be targeted, and protective factors promoted, to reduce the detrimental effect of COVID-19 and other pandemics on HCW mental health.

## Introduction

The rapid transmission rates and clinical severity of COVID-19 on patient health have brought global national health systems and their healthcare workers (HCWs) under considerable pressure. Whilst HCWs already experience high levels of job-related stress (1) and are at risk of poor psychological wellbeing (2), their highly-demanding work (3) will be exacerbated during a pandemic increasing risk of ‘burnout’ (4), poorer quality of care (5) and risk of developing other mental health problems (2). During epidemics, it has been shown that worse HCW mental health is associated with contact with infected people; redeployment; inadequate training; existing mental health disorders (6, 7) and ‘moral injury’ (distress from being unable to provide treatment) (8); while better support, protective equipment, clear communication (6, 9) and resilience (10) may protect mental health. The COVID-19 pandemic presents additional novel and specific challenges and risks to HCW mental wellbeing as they carry out their roles and responsibilities.

An initial study conducted in China early on in the COVID-19 pandemic found that HCW working during COVID-19 experienced a high prevalence of severe depression, anxiety and PTSD; and those were female, young, ‘frontline’, and working in Wuhan, were most at risk of these symptoms (11). Since then, COVID-19 has also had a profound effect on the UK health system, however, research to date has not addressed the impact on UK HCW wellbeing, warranting urgent attention. Identifying the risk factors associated with working during COVID-19 which are detrimental to mental health can provide targets by which their impact on HCW mental well-being may be mediated. This may, in turn, help maintain the efficacy of healthcare systems. Research to date has been in smaller HCW cohorts, outside the UK, and not included consideration of COVID-19-relevant risk factors, or only a limited range potential risk factors, which may affect HCW mental health. This study aimed to address these shortfalls and provide a comprehensive examination of the mental health of a large cohort of UK HCW and how it has been affected by the COVID-19 pandemic by: 1) quantifying the prevalence of high mental health symptoms in UK HCW shortly after the initial UK COVID-19 peak; 2) identifying factors associated with risk of, or protection from, these symptoms; also 3) quantifying how mental health changed compared to before COVID-19; 4) quantifying HCW worries at the time; and 5) revealing whether frontline, London, and BAME HCW, and those making challenging moral/medical decisions, had worse mental health symptoms compared to their counterparts.

## Method

### Design

We report cross-sectional baseline data acquired shortly after the peak of the COVID-19 pandemic in the UK (between 22^nd^April and 10^th^ May 2020 inclusive (see supplementary materials S1)) from an on-going, survey-based, longitudinal cohort study. The authors assert that all procedures contributing to this work comply with the ethical standards of the relevant national and institutional committees on human experimentation and with the Helsinki Declaration of 1975, as revised in 2008. All procedures involving human subjects/patients were approved by the University of Roehampton Ethics Committee (REF: PSYCH 20/361) and the UK Health Research Authority.

### Participants and survey dissemination

An online web-based survey, outlined below, was disseminated as follows (see also supplementary materials S2). All NHS R&D departments in the UK were contacted and asked to disseminate the survey synopsis and weblink to staff. Within the planned study period, 52 NHS services (see supplementary materials S2) agreed and promoted the study to staff either through direct circular emails, staff intranet, or both. The study synopsis survey link was also shared through clinical networks, social media and a study webpage. Lastly, text in the synopsis and survey encouraged respondents to share the survey link with other healthcare professionals. The study invitation text strongly encouraged all HCW to take part even if they did not feel affected by the impact of COVID-19. Eligible respondents were UK-based HCWs who were 18 years or older. Written informed consent was obtained from all subjects.

The survey was implemented on the Qualtrics platform and cross-sectional data on the following were collected:

1. Validated mental health scales measuring four symptom domains: The PHQ-9(12) (Patient Health Questionnaire) measures depressive symptoms in the last 2 weeks; the GAD-7(13) (General Anxiety Disorder-7) measures anxiety over the previous 2 weeks. The 22-item IES-R(14) (Impact of event scale – revised) measures PTSD symptoms over the past 7 days; and the PSS(15) (Perceived Stress Scale) measures perception of stress over the past month. Only individuals who had experienced a stressful or traumatic event *related to* COVID-19 were administered the IES-R. Also, the CD-RISC (Connor-Davidson Resilience Scale)(16) was administered which measures resilience.
2. Questions addressing potential risk and protective factors (see supplementary materials S3) of mental health symptoms were identified using a knowledge-based approach built on scientific literature, through focus groups, study team meetings, and survey piloting feedback. The items included could be clustered within the following themes: (a) Demographics and roles including working on the ‘frontline’ (directly engaged in diagnosing, treating, or caring for patients); (b) Workplace readiness and preparation; (c) Risk management including Personal Protective Equipment (PPE); (d) Experience of traumatic and stressful events; (e) Protective: being able to share stress at work.
3. Respondents also quantified their level of current worry on items concerning their work, personal lives and COVID-19 using a 10-point Likert scale (see supplementary S3 section G and table 6).
4. Additionally, statistical comparison of ratings (on a 5-point Likert scale) made *during* COVID-19 (at survey completion) and retrospectively in relation to wellbeing, worries and views about work *before* COVID-19, were calculated. These items were again selected from focus groups, study team meetings, and survey piloting feedback and included anxiety, depression, and stress items (see supplementary materials S3 (section F) and table 5).

### Data analysis

Analyses were conducted to (i) determine the prevalence of high mental health symptoms, (ii) reveal the risk and protective factors of having high levels of mental health symptoms, and (iii) quantify change in mental health from before COVID, and to investigate group differences in mental health symptoms.

### Prevalence of high mental health symptoms

‘High symptoms’ of depression and anxiety were determined by individuals scoring ≥10 on the PHQ and GAD scales (‘moderate’ and ‘severe’ symptoms). Severe stress was classified by a PSS score ≥24 (upper quartile) and an IES-R score of ≥26 was used to classify high PTSD symptoms.

### Risk and protective factors

The relationship between the potential risk and protective factors with prevalence of ‘high’ levels of each symptom were determined using chi-square analyses. Stepwise multivariable logistic regression analyses for each symptom domain (‘high symptoms’ vs not) were then performed and included the factors that were significant in chi-square analyses (0.05 significance level to enter / stay in the model).

### Change in mental health and group differences

Repeated-measures ANOVA were used to quantify (i) change in wellbeing from *pre* to *during* COVID-19 for the whole cohort, and to examine between-group differences between FL and NFL; (ii) to compare level of current worries between FL and NFL. Effect sizes were considered small=0.01, medium=0.06, large=0.14(17). Sensitivity analyses (ANOVA) revealed whether FL (vs NFL), BAME (vs non-BAME) workers, and those making challenging medical decisions (vs not) had higher mental health symptoms than their counterparts. Analyses were conducted in SAS 9.4 (SAS Institute Inc.) and SPSS v25 (IBM Corp., Armonk, N.Y., USA).

## Results

### Participant demographics and roles

3379 participants consented. Non-HCW and those who completed <70% of the survey were excluded leaving 2773 respondents (see table 1 for main descriptive statistics and tables S4-7).

**Table 1.** showing frequency (N, %) of demographics, roles, settings and COVID-19 status for the whole cohort, and stratified X^2^ (p) statistics for frontline (FL) vs non-frontline (NFL) and inside London vs outside London

|  | Total N (%) | FL N (%) | NFL (%) | p | London N (%) | Outside N (%) | p |
| --- | --- | --- | --- | --- | --- | --- | --- |
| **Sex** |  |  |  | 0.1905 |  |  | 0.0002 |
| Male | 395 (14.24%) | 188 (15.36%) | 166 (13.55%) |  | 135 (18.88%) | 259 (12.65%) |  |
| Female | 2365 (85.29%) | 1029 (84.07%) | 1056 (86.20%) |  | 577 (80.70%) | 1781 (86.96%) |  |
| Other / not reported | 13 (0.47%) | 7 (0.57%) | 3 (0.24%) |  | 3 (0.42%) | 8 (0.39%) |  |
| **Age categories** |  |  |  | <.0001 |  |  | 0.0002 |
| Under 25 | 102 (3.68%) | 61 (4.98%) | 29 (2.37%) |  | 18 (2.52%) | 83 (4.05%) |  |
| 25-34 | 630 (22.72%) | 301 (24.59%) | 264 (21.55%) |  | 207 (28.95%) | 420 (20.51%) |  |
| 35-44 | 680 (24.52%) | 306 (25.00%) | 306 (24.98%) |  | 172 (24.06%) | 508 (24.8%) |  |
| 45-54 | 813 (29.32%) | 374 (30.56%) | 345 (28.16%) |  | 186 (26.01%) | 625 (30.52%) |  |
| 55-64 | 508 (18.32%) | 174 (14.22%) | 259 (21.14%) |  | 120 (16.78%) | 386 (18.85%) |  |
| 65 and above | 36 (1.30%) | 7 (0.57%) | 21 (1.71%) |  | 12 (1.68%) | 24 (1.17%) |  |
| Not reported | 4 (0.14%) | 1 (0.08%) | 1 (0.08%) |  | 0 (0.00%) | 2 (0.01%) |  |
| **Ethnicity** |  |  |  | 0.0034 |  |  | <.0001 |
| White | 2414 (87.05%) | 1040 (84.97%) | 1094 (89.31%) |  | 546 (76.36%) | 1861 (90.87%) |  |
| Mixed / Multiple ethnic groups | 67 (2.42%) | 29 (2.37%) | 27 (2.20%) |  | 23 (3.22%) | 44 (2.15%) |  |
| Asian / Asian British | 191 (6.89%) | 107 (8.74%) | 62 (5.06%) |  | 78 (10.91%) | 113 (5.52%) |  |
| Black / African / Caribbean / Black British | 65 (2.34%) | 29 (2.37%) | 33 (2.69%) |  | 49 (6.85%) | 15 (0.73%) |  |
| Other | 19 (0.69%) | 10 (0.82%) | 6 (0.49%) |  | 12 (1.68%) | 7 (0.34%) |  |
| Not reported | 17 (0.61%) | 9 (0.74%) | 3 (0.24%) |  | 7 (0.98%) | 8 (0.39%) |  |
| **BAME** | 342 (12.33%) | 175 (14.30%) | 128 (10.45%) | 0.0029 | 162 (22.66%) | 179 (8.74%) | <.0001 |
| **Relationship** |  |  |  | 0.7375 |  |  | <.0001 |
| Single | 371 (13.38%) | 167 (13.64%) | 163 (13.31%) |  | 139 (19.44%) | 230 (11.23%) |  |
| In a relationship/married | 2209 (79.66%) | 973 (79.49%) | 977 (79.76%) |  | 526 (73.57%) | 1677 (81.88%) |  |
| Separated/divorced | 134 (4.83%) | 58 (4.74%) | 59 (4.82%) |  | 29 (4.06%) | 105 (5.13%) |  |
| Other | 28 (1.01%) | 15 (1.23%) | 10 (0.82%) |  | 3 (0.42%) | 25 (1.22%) |  |
| Not reported | 31 (1.12%) | 11 (0.90%) | 16 (1.31%) |  | 18 (2.52%) | 11 (0.54%) |  |
| **Work setting** |  |  |  | <.0001 |  |  | <.0001 |
| NHS – Hospital | 1411 (50.88%) | 790 (64.54%) | 456 (37.22%) |  | 330 (46.15%) | 1075 (52.49%) |  |
| NHS - GP surgery | 136 (4.90%) | 65 (5.31%) | 53 (4.33%) |  | 18 (2.52%) | 118 (5.76%) |  |
| NHS - Other community team | 590 (21.28%) | 184 (15.03%) | 344 (28.08%) |  | 134 (18.74%) | 455 (22.22%) |  |
| NHS - Mental Health trust | 393 (14.17%) | 81 (6.62%) | 276 (22.53%) |  | 185 (25.87%) | 207 (10.11%) |  |
| NHS - Ambulance service | 6 (0.22%) | 2 (0.16%) | 1 (0.08%) |  | 1 (0.14%) | 5 (0.24%) |  |
| A private hospital | 15 (0.54%) | 7 (0.57%) | 5 (0.41%) |  | 4 (0.56%) | 11 (0.54%) |  |
| Care home* | 45 (1.62%) | 28 (2.29%) | 14 (1.14%) |  | 7 (0.98%) | 38 (1.86%) |  |
| Nursing home* | 21 (0.76%) | 17 (1.39%) | 2 (0.16%) |  | 3 (0.42%) | 18 (0.88%) |  |
| Other (10) | 152 (5.48%) | 49 (4%) | 73 (5.96%) |  | 33 (4.62%) | 119 (5.81%) |  |
| Not reported | 4 (0.14%) | 1 (0.08%) | 1 (0.08%) |  | 0 (0.00%) | 2 (0.1%) |  |
| **Role** |  |  |  | <.0001 |  |  | <.0001 |
| Nurses/Midwife | 852 (30.72%) | 461 (37.66%) | 285 (23.27%) |  | 146 (20.42%) | 706 (34.47%) |  |
| Doctor | 386 (13.92%) | 229 (18.71%) | 112 (9.14%) |  | 148 (20.7%) | 237 (11.57%) |  |
| Allied Healthcare Professionals | 772 (27.84%) | 264 (21.57%) | 425 (34.69%) |  | 235 (32.87%) | 536 (26.17%) |  |
| Management | 245 (8.83%) | 39 (3.19%) | 174 (14.20%) |  | 85 (11.89%) | 158 (7.71%) |  |
| Other health worker | 499 (17.99%) | 206 (16.83%) | 220 (17.96%) |  | 98 (13.71%) | 379 (18.15%) |  |
| Not reported | 19 (0.69%) | 25 (2.04%) | 9 (0.73%) |  | 3 (0.42%) | 32 (1.56%) |  |
| **Mental health diagnosis (any) †** | 473 (17.06%) | 194 (15.85%) | 213 (17.39%) | 0.4378 | 101 (14.13%) | 372 (18.16%) | 0.0681 |
| **COVID-19 status** |  |  |  | <.0001 |  |  | 0.227 |
| Past positive | 80 (2.88%) | 56 (4.58%) | 19 (1.55%) |  | 19 (2.66%) | 61 (2.98%) |  |
| Past negative | 158 (5.7%) | 99 (8.09%) | 45 (3.67%) |  | 42 (5.87%) | 116 (5.66%) |  |
| Past - suspected | 574 (20.7%) | 257 (21%) | 239 (19.51%) |  | 181 (25.31%) | 392 (19.14%) |  |
| Current symptoms, tested, awaiting results | 21 (0.76%) | 17 (1.39%) | 3 (0.24%) |  | 8 (1.12%) | 12 (0.59%) |  |
| Current symptoms, not tested | 14 (0.5%) | 10 (0.82%) | 3 (0.24%) |  | 3 (0.42%) | 11 (0.54%) |  |
| No symptoms | 1784 (64.33%) | 719 (58.74%) | 876 (71.51%) |  | 425 (59.44%) | 1351 (65.97%) |  |
| Other | 142 (5.12%) | 66 (5.39%) | 40 (3.27%) |  | 37 (5.17%) | 105 (5.13%) |  |
| **Tested for COVID-19** |  |  |  | <.0001 |  |  | 0.227 |
| No | 2377 (85.72%) | 974 (79.58%) | 1117 (91.18%) |  | 620 (86.71%) | 1748 (85.35%) |  |
| Yes | 381 (13.74%) | 245 (20.02%) | 100 (8.16%) |  | 94 (13.15%) | 287 (14.01%) |  |
| Not reported | 15 (0.54%) | 5 (0.41%) | 8 (0.65%) |  | 1 (0.14%) | 13 (0.63%) |  |

*\* removed from main analyses due to low N* *∅ see supplementary tables for further breakdown* *BAME=Black Asian and Minority Ethnic*

### Prevalence of high mental health symptoms

Table 2 shows cohort mental health symptoms. 28.1% (374) had high depression, 33.1% (919) high anxiety; and 27.5% (750) were in the top quartile for stress (see table). 60.6% (1681) had experienced a stressful or traumatic event related to COVID-19 and 14.6% (404) reported high PTSD symptoms.

**Table 2.** showing frequency distributions (N (%)) and median/means/IQR for each symptom scale, for the total cohort and stratified X^2^ (p) statistics for frontline (FL) vs non-frontline (NFL) and inside London vs outside London

|  | Total | Total % | FL | % | NFL | % | p | Inside London | % | Outside London N | % | p |
| --- | --- | --- | --- | --- | --- | --- | --- | --- | --- | --- | --- | --- |
| **PHQ median (IQR)** | 6 (7) |  | 7 (9) |  | 5 (7) |  |  | 5 (7) |  | 6 (8) |  |  |
| **PHQ mean (sd)** | 7.18 (6.07) |  | 8.00 (6.21) |  | 6.97 (5.75) |  |  | 6.53 (5.58) |  | 7.41 (6.22) |  |  |
| Normal (0-5) | 1172 | 42.26 | 475 | 38.81 | 582 | 47.51 | <.0001 | 330 | 46.15 | 837 | 40.87 | 0.0064 |
| Mild (5-9) | 823 | 29.68 | 369 | 30.15 | 343 | 28 |  | 217 | 30.35 | 605 | 29.54 |  |
| Moderate (10-14) | 404 | 14.57 | 198 | 16.18 | 155 | 12.65 |  | 93 | 13.01 | 309 | 15.09 |  |
| Moderately severe (15-19) | 231 | 8.33 | 104 | 8.5 | 98 | 8 |  | 53 | 7.41 | 176 | 8.59 |  |
| Severe (20-27) | 143 | 5.16 | 78 | 6.37 | 47 | 3.84 |  | 22 | 3.08 | 121 | 5.91 |  |
| **GAD median (IQR)** | 6 (8) |  | 8 (9) |  | 6 (8) |  |  | 6 (7) |  | 7 (9) |  |  |
| **GAD mean (sd)** | 7.51 (5.58) |  | 8.71 (5.83) |  | 7.34 (5.14) |  |  | 6.86 (5.19) |  | 7.72 (5.70) |  |  |
| Normal (0-4) | 1014 | 36.57 | 376 | 30.72 | 524 | 42.78 | <.0001 | 288 | 40.28 | 723 | 35.3 | 0.0004 |
| Mild (5-9) | 840 | 30.29 | 375 | 30.64 | 369 | 30.12 |  | 235 | 32.87 | 602 | 29.39 |  |
| Moderate (10-14) | 532 | 19.18 | 254 | 20.75 | 211 | 17.22 |  | 118 | 16.5 | 411 | 20.07 |  |
| Severe ($\geq 15$) | 387 | 13.96 | 219 | 17.89 | 121 | 9.88 | | 74 | 10.35 | 312 | 15.23 | |
| **IES-R median (IQR)\*** | 16 (23) |  | 18 (25) |  | 13 (17) |  |  | 15 (22) |  | 16 (23) |  |  |
| **IES-R mean (sd)** | 20.30 (16.68) |  | 22.05 (17.1) |  | 16.4 (14.6) |  |  | 19.89 (16.57) |  | 20.43 (16.73) |  |  |
| Q1 < 7.00 | 387 | 13.96 | 202 | 16.5 | 140 | 11.43 | <.0001 | 104 | 14.55 | 282 | 13.77 | 0.9762 |
| Q2 7.00 ≤ IESR < 20.30 | 620 | 22.36 | 340 | 27.78 | 203 | 16.57 | | 162 | 22.66 | 456 | 22.27 | |
| Q3 20.30 ≤ IESR < 30.00 | 248 | 8.94 | 154 | 12.58 | 65 | 5.31 | | 63 | 8.81 | 185 | 9.03 | |
| Q4 $\geq 30.00$ | 426 | 15.36 | 294 | 24.02 | 85 | 6.94 | | 106 | 14.83 | 318 | 15.53 | |
| **CDRISC median (IQR)** | 28 (9) |  | 28 (9) |  | 27 (9) |  |  | 27 (9) |  | 28 (9) |  |  |
| **CDRISC mean (sd)** | 27.5 (6.48) |  | 27.9 (6.43) |  | 27.5 (6.34) |  |  | 27.36 (6.53) |  | 27.49 (6.47) |  |  |
| Q1 < 23.00 | 567 | 20.45 | 244 | 19.93 | 262 | 21.39 | 0.0173 | 149 | 20.84 | 416 | 20.31 | 0.2268 |
| Q2 23.00 <= <27.46 | 720 | 25.96 | 289 | 23.61 | 349 | 28.49 |  | 205 | 28.67 | 514 | 25.1 |  |
| Q3 27.46 <= < 32.00 | 633 | 22.83 | 289 | 23.61 | 264 | 21.55 |  | 152 | 21.26 | 478 | 23.34 |  |
| Q4 => 32.00 | 688 | 24.81 | 318 | 25.98 | 287 | 23.43 |  | 175 | 24.48 | 513 | 25.05 |  |
| **PSS median (IQR)** | 19 (11) |  | 20 (10) |  | 19 (10) |  |  | 18 (11) |  | 19 (10) |  |  |
| **PSS mean (sd)** | 18.6 (7.47) |  | 19.6 (7.45) |  | 18.6 (6.93) |  |  | 18.06 (7.47) |  | 18.73 (7.48) |  |  |
| Q1 <11 | 608 | 21.93 | 233 | 19.04 | 319 | 26.04 | <.0001 | 173 | 24.2 | 433 | 21.14 | 0.123 |
| >11 Q2 <13 | 727 | 26.22 | 303 | 24.75 | 328 | 26.78 |  | 198 | 27.69 | 525 | 25.63 |  |
| >13 Q3 <24 | 688 | 24.81 | 318 | 25.98 | 298 | 24.33 |  | 165 | 23.08 | 520 | 25.39 |  |
| Q4 >24 | 750 | 27.50 | 370 | 30.23 | 280 | 22.85 |  | 179 | 25.03 | 570 | 27.83 |  |

*GAD= General Anxiety Disorder-7; IES-R= Impact of event scale – revised; PHQ=Patient Health Questionnaire; PSS= Perceived Stress Scale.*
*CD-RISC=Connor-Davidson Resilience Scale*
*\* 60.5% of the sample completed IES-R – those who identified experiencing a stressful or traumatic event related to COVID-19.*

### Predictive models of high mental health symptoms

All stepwise multivariate logistic regression models converged with no significant collinearity of factors, or residual data due to missingness. All models were highly significant (likelihood ratio, core, and Wald p<.0001) presenting good fit and high prediction capabilities (c score = 0.739 – 0.82). Significant factors retained in each model of symptoms with odds ratios are shown in table 3. Frequency distributions are shown in table 4; chi-square summaries are reported in supplementary materials S8.

**Table 3.**
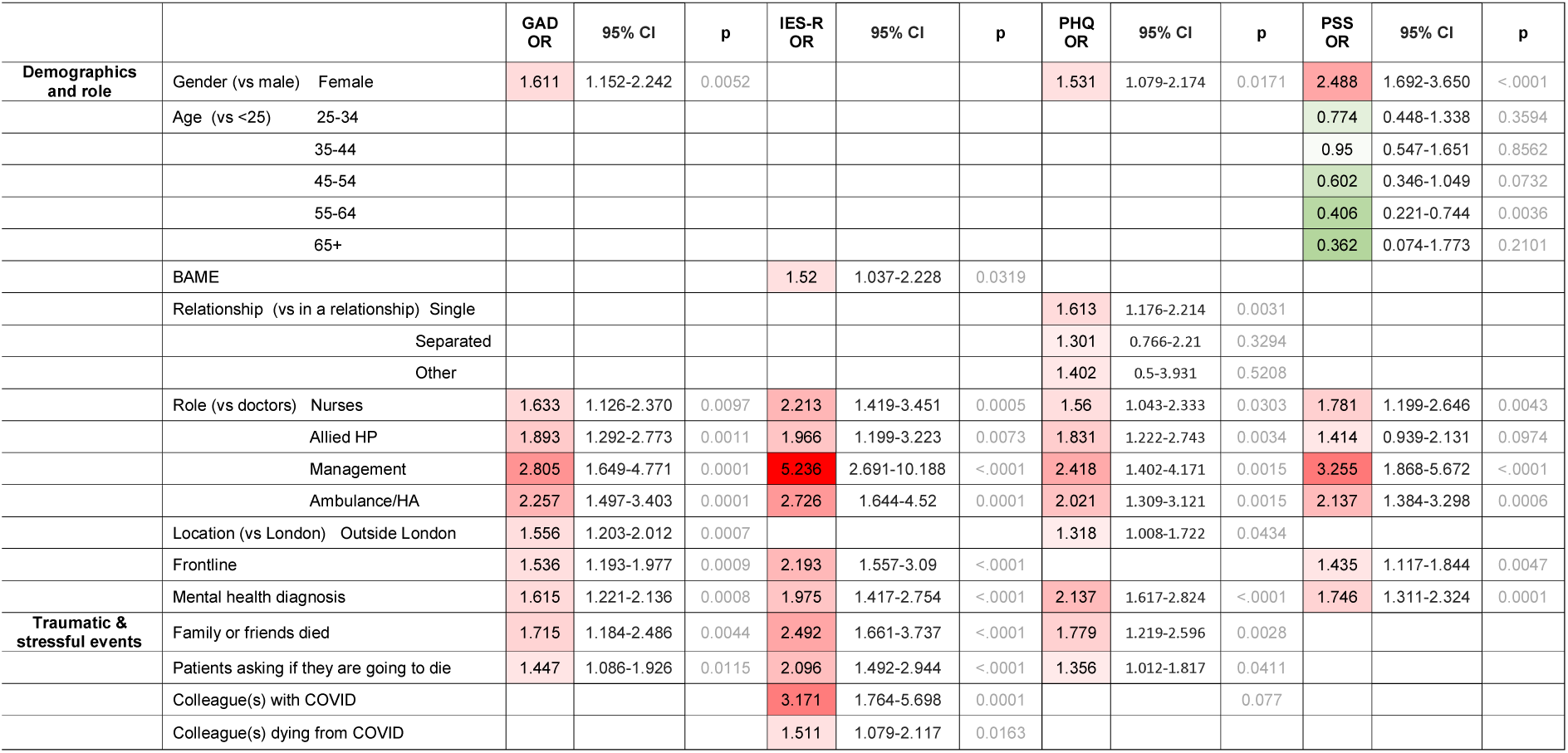
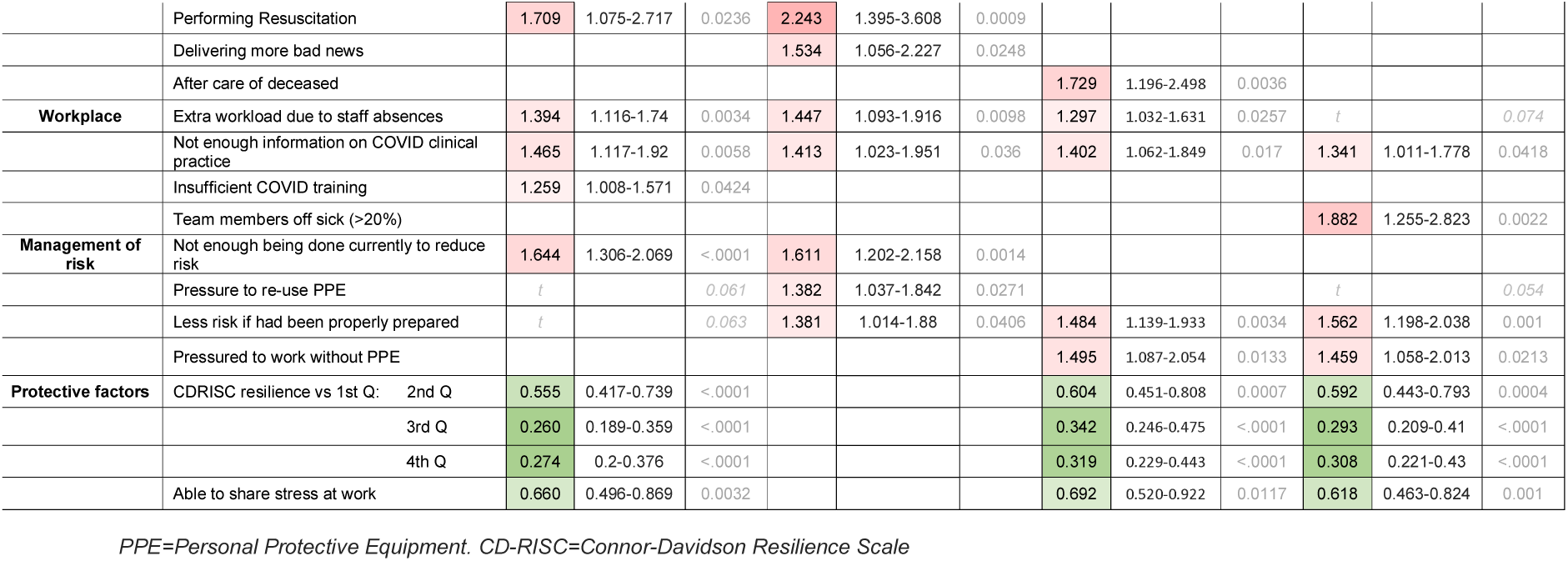
summary table showing odds ratios (OR) and p values for risk and protective factors retained in each model. Red shading = risk, green shading = protection; darker shading reflects stronger effects. Blank cells show the factor was not significantly retained in the model of the outcome score. “t” refers to trend-level effects shown for illustration purposes only.

**Table 4.** showing frequency distributions (N(%) (the percentage of the symptoms groups (columns) constituted by the ‘factor’ group) of 2773 total sample for high and low levels of mental health symptoms for all significant variables in the logistic regressions for the four scales (reported in table 3).

|  |  | **GAD low (N**<br>**(%))** | **GAD high**<br>**(N (%))** | **IES-R low**<br>**(N (%))** | **IES-R high**<br>**(N (%))** | **PHQ low**<br>**(N (%))** | **PHQ high**<br>**(N (%))** | **PSS low**<br>**(N (%))** | **PSS high**<br>**(N (%))** |
| --- | --- | --- | --- | --- | --- | --- | --- | --- | --- |
| **Demographics**<br>**and role** | Gender: Female | 1547 (83.4) | 818 (89.0) | 2049 (85.4) | 316 (87.8) | 1678 (84.4) | 687 (88.9) | 1907 (84.4) | 458 (91.6) |
|  | Age: Under 25 | 55 (3.0) | 47 (5.1) | 79 (3.3) | 23 (6.4) | 66 (3.3) | 36 (4.6) | 72 (3.2) | 30 (6) |
|  | 25-34 | 381 (20.6) | 249 (27.2) | 529 (22) | 101 (27.9) | 429 (21.5) | 201 (25.9) | 474 (20.9) | 156 (31.1) |
|  | 35-44 | 434 (23.4) | 246 (26.8) | 592 (24.6) | 88 (24.3) | 493 (24.7) | 187 (24.1) | 538 (23.7) | 142 (28.3) |
|  | 45-54 | 573 (30.9) | 240 (26.2) | 715 (29.7) | 98 (27.1) | 593 (29.8) | 220 (28.4) | 704 (31) | 109 (21.8) |
|  | 55-64 | 380 (20.5) | 128 (14.0) | 457 (19) | 51 (14.1) | 382 (19.2) | 126 (16.2) | 446 (19.6) | 62 (12.4) |
|  | 65+ | 29 (1.6) | 7 (0.8) | 35 (1.5) | 1 (0.3) | 30 (1.5) | 6 (0.8) | 34 (1.5) | 2 (0.4) |
|  | BAME | 224 (12.1) | 118 (13.0) | 284 (11.9) | 58 (16.0) | 272 (12.0) | 106 (13.7) | 272 (12.0) | 70 (14.1) |
|  | Single | 227 (12.4) | 144 (15.9) | 312 (13.1) | 59 (16.4) | 235 (11.9) | 136 (17.7) | 300 (13.3) | 71 (14.4) |
|  | In a relationship | 1497 (81.6) | 712 (78.4) | 1931 (81) | 278 (77.4) | 1625 (82.3) | 584 (76) | 1813 (80.6) | 396 (80.5) |
|  | Separated/Divorced | 92 (5.0) | 42 (4.6) | 115 (4.8) | 19 (5.3) | 96 (4.9) | 38 (4.9) | 114 (5.1) | 20 (4.1) |
|  | Other | 18 (1.0) | 10 (1.1) | 25 (1.0) | 3 (0.8) | 18 (0.9) | 10 (1.3) | 23 (1.0) | 5 (1.0) |
|  | Nurses | 560 (30.4) | 292 (32.0) | 707 (29.5) | 145 (40.2) | 611 (30.9) | 241 (31.1) | 705 (31.3) | 147 (29.5) |
|  | Doctors | 300 (16.3) | 86 (9.4) | 346 (14.5) | 40 (11.1) | 318 (16.1) | 68 (8.8) | 328 (14.5) | 58 (11.6) |
|  | Allied Healthcare | 528 (28.7) | 244 (26.7) | 704 (29.4) | 68 (18.8) | 564 (28.5) | 208 (26.9) | 641 (28.4) | 131 (26.3) |
|  | Management | 162 (8.8) | 83 (9.1) | 216 (9) | 29 (8.0) | 175 (8.8) | 70 (9.0) | 198 (8.8) | 47 (9.4) |
|  | Ambulance/HA | 291 (15.8) | 208 (22.8) | 420 (17.6) | 79 (21.9) | 312 (15.8) | 187 (24.2) | 383 (17) | 116 (23.2) |
|  | Location Outside London | 1325 (71.7) | 723 (79.0) | 1777 (72.0) | 271 (75.1) | 1442 (72.5) | 606 (78.3) | 1667 (73.7) | 381 (76.2) |
|  | Frontline | 751 (45.6) | 473 (58.8) | 966 (45.5) | 258 (79.1) | 844 (47.7) | 380 (55.9) | 973 (48.2) | 251 (58.5) |
|  | Non-Frontline | 893 (54.3) | 332 (41.2) | 1157 (54.0) | 68 (20.8) | 926 (52.2) | 300 (44.1) | 1047 (51.2) | 178 (41.5) |
|  | Mental health diagnosis | 238 (13.1) | 235 (26.2) | 377 (15.9) | 96 (27.1) | 244 (12.4) | 229 (30.2) | 325 (14.5) | 148 (30.6) |
| **Traumatic & stressful events** | Family or friends died | 132 (7.2) | 112 (12.6) | 173 (7.3) | 71 (20.4) | 143 (7.3) | 101 (13.5) | 185 (8.3) | 59 (12.2) |
|  | Patients asking if they are going to die | 329 (17.7) | 256 (27.9) | 419 (17.4) | 166 (45.7) | 375 (18.8) | 210 (27.0) | 437 (19.2) | 148 (29.5) |
|  | Colleague(s) with COVID-19 | 1457 (78.6) | 776 (84.4) | 1894 (78.6) | 339 (93.4) | 1569 (78.6) | 664 (85.3) | 1797 (79.1) | 436 (87.0) |
|  | Colleague(s) dying from COVID-19 | 238 (12.8) | 157 (17.1) | 303 (12.6) | 92 (25.3) | 276 (13.8) | 119 (15.3) | 313 (13.8) | 82 (16.4) |
|  | Performing Resuscitation | 70 (3.8) | 64 (7.0) | 78 (3.2) | 56 (15.4) | 89 (4.5) | 45 (5.8) | 104 (4.6) | 30 (6.0) |
|  | Delivering more bad news than usual | 249 (13.4) | 172 (18.7) | 290 (12.0) | 131 (36.1) | 278 (13.9) | 143 (18.4) | 323 (14.2) | 98 (19.6) |
|  | After care of deceased | 147 (7.9) | 135 (14.7) | 187 (7.8) | 95 (26.2) | 169 (8.5) | 113 (4.5) | 208 (9.2) | 74 (14.8) |
| **Workplace** | Extra workload due to staff absences | 868 (48.0) | 520 (58.0) | 1139 (48.5) | 249 (70.0) | 944 (48.5) | 444 (58.5) | 1092 (49.3) | 296 (60.2) |
|  | Not enough information on COVID-19 clinical practice | 276 (15.0) | 283 (31.0) | 418 (17.4) | 141 (39.3) | 323 (16.3) | 236 (30.5) | 396 (17.5) | 163 (32.6) |
|  | Insufficient COVID-19 training | 775 (42.1) | 448 (49.1) | 1056 (44.1) | 167 (46.3) | 833 (42.1) | 390 (50.4) | 968 (43.0) | 255 (50.9) |
|  | Team members off sick (>20%) | 212 (11.4) | 179 (19.4) | 288 (11.9) | 103 (28.4) | 229 (11.5) | 162 (20.8) | 270 (11.9) | 121 (24.2) |
| **Management of risk** | Not enough being done currently to reduce risk | 526 (28.5) | 448 (49.1) | 776 (32.4) | 198 (55.0) | 612 (30.8) | 362 (46.9) | 726 (32.3) | 248 (49.7) |
|  | Pressure to re-use PPE | 397 (22.6) | 316 (36.2) | 546 (23.9) | 167 (49.0) | 462 (24.3) | 251 (34.3) | 539 (25.0) | 174 (36.5) |
|  | Less risk if had been properly prepared | 601 (41.4) | 309 (51.6) | 805 (42.5) | 105 (52.5) | 654 (42.4) | 256 (50.2) | 765 (43.8) | 145 (47.2) |
|  | Pressured to work without PPE | 182 (10.5) | 180 (21.0) | 268 (11.9) | 94 (28.2) | 209 (11.1) | 153 (21.5) | 251 (11.8) | 111 (23.7) |
| **Protective factors** | CDRISC resilience 1st Q: | 266 (15.2) | 301 (34.9) | 461 (20.3) | 106 (31.9) | 311 (16.5) | 256 (35.2) | 372 (17.4) | 195 (41) |
|  | 2nd Q | 463 (26.5) | 257 (29.8) | 636 (27.9) | 84 (25.3) | 510 (27.1) | 210 (28.9) | 580 (27.2) | 140 (29.4) |
|  | 3rd Q | 477 (27.3) | 156 (18.1) | 568 (25) | 65 (19.6) | 498 (26.5) | 135 (18.6) | 564 (26.5) | 69 (14.5) |
|  | 4th Q | 539 (30.9) | 149 (17.3) | 611 (26.8) | 77 (23.2) | 562 (29.9) | 126 (17.3) | 616 (28.9) | 72 (15.1) |
|  | Able to share stresses at work | 1588 (87.8) | 673 (75.4) | 2010 (85.1) | 251 (73.0) | 1700 (86.8) | 561 (75.2) | 1909 (86.2) | 352 (72.0) |

### Anxiety

As shown in table 3, high anxiety was significantly associated with being female, all non-doctor roles (vs doctor), working outside London, being frontline, and having a mental health diagnosis. Friends or family dying from COVID-19, patients asking if they are going to die, and performing resuscitation were also associated with high anxiety, as were insufficient training, extra workload, insufficient information and thinking not enough is currently being done to reduce risk.

### PTSD

All non-doctor (vs doctor) roles - particularly being a manager, and being FL, having BAME status, and existing mental health conditions significantly raised the risk of high PTSD symptoms, as did experience of all traumatic and stressful events except aftercare of the deceased. High PTSD symptoms were also significantly associated with pressure to re-use PPE; insufficient information; perception that not enough had been, nor was being done, to reduce risk; and greater workload.

### Depression

High depression was significantly associated with being female, all non-doctor roles (vs doctor), working outside London, and having a mental health diagnosis. Those experiencing friends or family dying, patients asking them if they are going to die, and performing aftercare for the deceased had greater risk. Extra workload, pressure to work without PPE, insufficient information and perception that not enough had been done to reduce risk also significantly raised risk of high depression.

### Stress

Being female, older (55-64 years vs <25), all non-doctor roles (vs doctor), working on the frontline, and having a mental health diagnosis were associated with significantly increased likelihood of high stress; as were insufficient information, pressure to work without PPE, >20% of team members off sick, and perception that not enough had been done to reduce risk were also significantly associated with high symptoms.

### Protective factors

Greater resilience and being able to share stress at work were associated with significantly lower likelihood of high anxiety, stress and depression though not PTSD.

### Change from pre-COVID-19 to during COVID-19

All items, including stress, mood and anxiety, were rated as significantly worse during COVID-19 compared to pre-COVID-19, many to a highly significant level with very high effect sizes (see supplementary table S9). Being worried about family health showed the greatest (negative) change. FLs made similar pre-COVID-19 ratings to NFLs across all factors but reported significantly greater worsening for most items including stress, depression and anxiety (illustrated in supplementary figure S11).

### Frontline, London and BAME workers

FLs were significantly more likely to be more depressed, anxious, have high PTSD symptoms and be more stressed than NFL (all p<.0001). Working in London was associated with *lower* risk of depression (p<.01) and anxiety (p<.0005) than outside London (though no difference in stress or PTSD). BAME status (N=342) was significantly associated with greater risk of high PTSD symptoms (OR=1.52), but not high anxiety, stress, or depression.

Post-hoc analyses were conducted to explore BAME experiences further. Physically *‘at-risk’* BAME individuals (N=85) did not have higher mental health symptoms than BAME not *‘at-risk’* (N=257), nor compared to *non*-BAME individuals physically *‘at risk’* (N=593) (all ps>0.45). BAME individuals were, however, significantly more worried about contracting COVID-19 at work (mean(sd)=3.09(1.08) vs non-BAME mean(sd) = 2.67(1.07); t(2754)=6.84, p<.0001); being uncertain of having COVID-19 (mean(sd)=2.77(1.16) vs non-BAME mean(sd)=2.21(1.09); t(2754)=8.93, p<.001); getting ill or dying from COVID-19 (mean(sd)=2.2(1.12) vs non-BAME mean(sd)=1.88 (1.19), t(2754)=4.74, p<.001) and lack of PPE (mean(sd)=2.86 (1.14) vs non-BAME mean(sd) = 2.39(1.12), t(2754)=7.29, p<.001).

### Medical decision-making

11.1% (307) of respondents were in a position to make decisions about whether patients received treatment and 17.9% (53) had denied treatment to a patient (see supplementary materials S7). They reported significantly more anxiety (but not depression, PTSD (though strong trend), or stress (though strong trend)) than those (N=39) who had not denied treatment (GAD mean=8.00 (5.94) vs 5.46 (5.19), t(90)=2.13, p<.05). Those with support of an ethics panel in decision-making (58.0% (178)) were significantly less stressed (PSS mean=16.98 (8.1) vs mean=19.06 (7.33), t(302)=2.29, p<.01) but were not significantly less depressed, anxious, nor had lower PTSD symptoms than those without support (N=126).

### Worries

Across the cohort, worry was greatest for family and friends becoming ill or dying from COVID-19 followed by worries that they will infect them (see supplementary table S10); and lowest for their own mental health and about poor workplace management. FL were significantly more worried than NFL for all concerns (all p<.001 except ‘ability to support others’ p<0.05).

## Discussion

To our knowledge this is the first study examining the impact of COVID-19 on the mental health of HCWs in the UK. A significant proportion reported high depression (28%), high anxiety (33%), and high COVID-19-related PTSD symptoms (15%). Across the cohort, mental health indicators had significantly deteriorated compared to before COVID. Analyses revealed a set of *fixed* (demographic and role-related) and a separate set of *controllable* factors which were significantly associated with high levels of mental health symptoms in HCW.

The *fixed* risk factors for high mental health symptoms were being female, all roles compared to doctor, working on the ‘frontline’, and having an existing mental disorder. Being single was a risk for high depression and being younger to stress. While some of these components have been identified in previous pandemics(6, 9) and recent research of much smaller cohorts outside the UK(11, 18–20) the present study expands significantly on this work in several ways in terms of sample size, comprehensive examination of risk factors beyond demographics and roles, and scope of findings. We show, that allied HCW, and particularly managers, were at significantly increased risk of high symptoms. Managers, in particular, were 5.2 times more likely to report high PTSD symptoms - likely due to additional pressures and the rapid changes COVID-19 brings to their healthcare settings as well as increased threat to patients, staff and themselves. This suggests the need for additional support for personnel in these roles.

Importantly, a cluster of *controllable* risk factors relating to workplace characteristics and role-related activities were also significantly associated with high mental health symptoms. Pressure to work without PPE, and that risk from COVID-19 could have been reduced with better workplace preparation, were associated with high depression. These factors were also associated with high stress along with practical issues such as absent team members and lack of sufficient information on clinical procedures. The effects of having additional workload were broad – being linked to high anxiety, depression and PTSD, while insufficient training was uniquely associated with high levels of anxiety (also shown after SARS((21)). High anxiety and PTSD were additionally associated with insufficient action being taken to reduce risk. There was a further critical role of a lack of sufficient information on COVID-19 clinical practice - being linked to high symptoms in all domains.

Critically then, a number of preventable workplace factors relating to perception of personal risk specifically increases the likelihood of high PTSD symptoms: pressure to re-use PPE and failure of the workplace to reduce risk through preparation. A strong link between risk perception and PTSD has been reported previously during SARS(21–25). As subjective appraisal of threat may contribute more to PTSD development than objective trauma severity (26) a sense of persistent danger to the self may catalyse the development of PTSD symptoms. This also highlights that perception of risk goes beyond PPE availability and includes multiple systemic and organisational components within healthcare settings. As a longer or repeated exposure (see)(21)(27) raises risk of PTSD, more adequate PPE and workplace preparation may mitigate future development of PTSD. It is indeed noteworthy that over half of respondents stated that more PPE would reduce their anxiety.

Unlike an earlier study from China(11), an epicentre effect was not apparent. Working in London was associated with *lower* risk of anxiety and depression. While London workers were significantly less female, less likely to be pressured to reuse PPE, or expect to get severely ill (each linked to lower risk), they *also* experienced a number of risk factors. These effects could be due to better-resourced healthcare settings, being more accustomed to stress from city living, or that Wuhan was the first global city experiencing a new, fatal virus.

Unsurprisingly, traumatic events predicted with high symptom scores, particularly PTSD. Personal loss and patients asking if they were going to die significantly raised the risk of high PTSD, anxiety and depression. Having colleagues with, or dying from, COVID-19 also significantly increased the likelihood of high PTSD symptoms but this was greater with respect to friends or family dying. A peer who contracts or dies from COVID may be more indicative of an on-going threat of danger to the self. Experiences where death is evident (patients dying and delivering bad news) also raised risk of PTSD symptoms (also seen following SARS)(22). Together, personal threat raised risk of high PTSD symptoms while the impaired readiness to work effectively in response to COVID-19 was linked to high anxiety - perhaps due to these being preventable. Lastly, aftercare for the deceased was uniquely linked to depression; performing resuscitation raised risk of high anxiety and PTSD; while practical issues increased risk of high stress.

Moral injury may contribute to the development of mental health symptoms(8). Here, HCWs who had denied treatment to patients were more anxious than those who had not, while support from an ethics panel was associated with lower stress highlighting the protective effects of shared decision-making on HCW mental wellbeing. The higher risk to managers may be due to such moral injury and the inability to adequately treat patients or protect staff.

Evidence that BAME individuals are at elevated physical risk of COVID-19 was first published near the survey start date(28). While BAME HCW were more likely to report high PTSD symptoms this was not accompanied by a significantly greater risk of high anxiety, depression or stress. Being ‘physically at high risk’ of COVID-19 was not associated with high mental health *symptoms*, but greater *worry* about self-protection. Elevated prevalence of PTSD in BAME individuals has previously been reported(29) and is associated with ‘additional life stress’(30). While more research on PTSD in BAME individuals should be done, this finding may reflect the same sense of sustained threat.

Across the cohort, wellbeing indicators broadly worsened during COVID-19 compared to before. FLs had significantly greater worries than NFL and were more also likely to be more depressed, anxious, and stressed than NFL - and were 2.1 times as likely to have high PTSD symptoms - likely due to the traumatic and stressful duties they perform, as well as their concerns about risk, PPE access and preparation.

Resilience and the ability to share stress at work were significantly protective from high symptoms except PTSD. Inadequate support has previously been shown to raise the risk of psychiatric morbidity in FL(6, 7, 22). Contrastingly, however, scores on the “*I need psychological help?*” item were low as were HCW worries about their ‘own mental health’. Staff may indeed prefer practical help such as more rest or PPE to psychological support(31). Resilience training may improve resistance to poor wellbeing, although this has been insufficiently researched in healthcare settings. PTSD risk may not be attenuated by resilience perhaps due to the more automatic and physiological, rather than cognitive, nature of these symptoms.

The study has several strengths and limitations. We recruited a large sample, near the peak of the COVID-19 UK outbreak, and the study provides the most comprehensive picture to date of the negative psychological impact of HCW to COVID-19 in the UK and associated risk factors. Participation in online surveys involves self-selection and respondents may not be fully representative. However, this approach permitted a rapid response around a critical period very close to the COVID-19 peak. The survey was de facto open to *all* HCW in the UK, and a significant proportion of all UK NHS sites (N=52) recruited staff to the study, providing very wide geographical coverage and a large sample size. That the sample characteristics are similar to the wider NHS workforce in terms of female:male ratio (85%, NHS=77%) and BAME proportion (13% vs NHS 19%) indicates that the data are broadly representative.

Pre-COVID-19 wellbeing scores derived from ratings which may not be fully accurate as they were retrospective, however, evidence suggests that ratings of past events in depressed individuals are reliable(32). If low mood resulted in more negative past ratings(33), this would only *increase* the effect sizes of worsening suggesting these effects are robust. Mood scores indicated that the cohort were not a *particularly* anxious or worrisome group per se and the majority of respondents reported only low or mild symptoms of anxiety and depression and low worry levels before COVID-19. Mental illness diagnosis frequency was also low and very similar to rates in the general population(29), while symptom scale scores and PTSD prevalence were similar to comparable studies(6, 11). Lastly, the study information also strongly encouraged those who felt they “were *not* be affected by COVID outbreak” to take part, so we would “have a complete view of HCW mental health” to prevent respondents assuming they should only complete if affected. The scales used were self-report and not diagnostic but have strong validity and reliability and are commonly used. This survey was cross-sectional but planned follow-up surveys will permit longitudinal analysis of effects and relationships. Finally, additional factors not examined may have a role in HCW mental health.

In conclusion, the COVID-19 pandemic has had a discernible and detrimental effect on the mental health and well-being of UK HCWs. High symptoms of poor mental health were prevalent, and markers of wellbeing had worsened compared to before COVID. A number of *fixed* factors raised risk of poor mental wellbeing, however, a separate set of *controllabl*e factors were identified that also raised risk - and these reflected elevated perception of COVID-19 risk and inadequate workplace preparedness. Critically, these findings can guide management strategy such as by improving PPE availability, training, communication of information, and management of staff absence. These are readily amenable targets and may reduce the risk of HCWs developing poor mental health during COVID-19, or other pandemics.

The study also strongly indicates that psychological risk assessments should be carried out based on the risk factors identified. All staff should be monitored for poor mental health and those showing high symptoms should be referred to mental health services. Employers should improve initiatives for HCWs to share stress particularly those with risk factors, and those making challenging treatment decisions. Bespoke interventions could be developed which target these risk factors, such as role- or duty-specific training. Improving resilience, perhaps through training, may also be effective. Finally, HCW show only low recognition of the importance of their own mental health so awareness of this should be raised. Lastly, working as a HCW during an pandemic can result in long term effects on mental health, which may persist for years(34). Attenuating these risks may help reduce the possibility of a major mental health crisis in UK healthcare and protect and retain HCWs. This is critical to delivery of effective treatment for patients and for planning a response to a second wave or future epidemic / pandemic - or in other countries where HCWs are yet to experience the impact on their mental health.

## Data Availability

We will engage with any reasonable request to share data.

## Funding Statement

This research received no specific grant from any funding agency, commercial or not-for-profit sectors. Use of scales and webhosting/advertising were covered by a small amount of charitable donations to the project from members of the public (∼£500).

### Acknowledgements

We first want to thank all the health care workers who shared their experiences with us. We also would like to thank those who contributed their time and hard work to the COVIDA Project: Natasha Ramachandran (UCL, London, UK); Carl Stratton for website design and media strategy; Eirini Koutsouroupa for media coordination. We also thank Mandy Holmes and Jan Harrison of the University of Roehampton for facilitating a rapid set-up of the study; and Dr. Lin Fou for proof-reading the manuscript.

## Author Contributions

JG was the study PI, study coordinator, and lead author. MF and JG led on the development and planning of the study and LV, AS, CS also contributed. AS conducted the Chi-Sq and regression analysis, assisted by MF and JG. JG conducted the remaining analyses. LV led on the survey dissemination strategy and liaising with NHS and non-NHS sites for study adoption. All authors worked on the development of the survey. JG is responsible for the overall content as guarantor.

## Data Availability

The data that support the findings of this study are available from the corresponding author, JG, upon reasonable request.

