## supplementary materials for "The impact of the COVID-19 pandemic on the mental health and wellbeing of UK healthcare workers" (from another version of this preprint: version 1, 2020-10-27)

**Contents**

S1: Figure showing survey period

S2: Survey dissemination.

S3: Covida Study Survey

S4: Table 1b showing additional demographics, health status and medications for the whole cohort, and stratified X^2^ (p) statistics for frontline (FL) vs non-frontline (NFL) and London vs outside London.

S5: Table 7 showing frequency (N (%)) distributions and X^2^ statistics (p) for the additional significant predictors retained in the symptom models (shown in table 3) for the total cohort and stratified X^2^ (p) statistics for frontline (FL) vs no

n-frontline (NFL) and inside London vs outside London

S6: Table 8 showing PPE and risk management factors for the whole cohort, and stratified X^2^ (p) statistics for frontline (FL) vs non-frontline (NFL) and London vs outside London.

S7: Table 9 showing traumatic and stressful events or duties for the whole cohort, and stratified X^2^ (p) statistics for frontline (FL) vs non-frontline (NFL) and London vs outside London

S8: Chi-sq summary statistics table (showing X*^2^* and p values) for factors and associated with each symptom outcome score. Shaded cells are p<.05.

S9: Table showing pre- and during-COVID-19 ratings for ‘negative’ and ‘positive’ factors (scored 1-5), and effect size of change (time). Also shown are group (Frontline/Non-frontline) x time (pre / during COVID-19) interaction effects (and description of which group showed the greatest change) from between-groups repeated-measures ANOVA.

S10: Table showing ratings of worry across a range of factors ordered by degree of worry for the total cohort as well as a statistical comparison of worries of frontline (FL) and non-frontline (NFL) workers (t, p).

S11: Figures showing change in low mood (left) and feeling anxious (centre) and feeling stressed (right), from pre to during COVID for frontline and non-frontline HCW. Error bars are 95% CI.

**S1: Figure showing the timing of the survey period with respect to the UK COVID-19 deaths.**

**The background plot on deaths by date is taken from the UK gov. Website (https://coronavirus.data.gov.uk/#category=nations&map=case).**

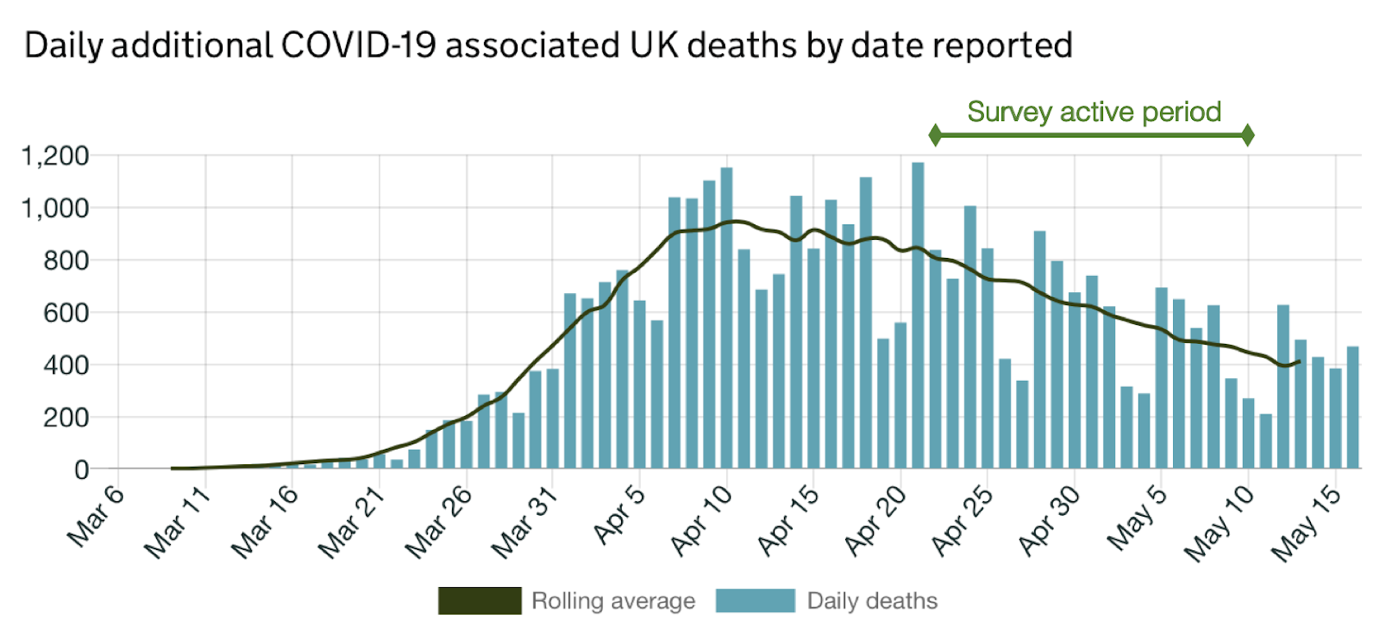

**S2 Survey dissemination**

We made contact with the all UK NHS research and development departments that had an email address publicly available. These were in total 262 NHS services R&D departments within the UK:

- 114 NHS foundations trusts
- 95 NHS trusts
- 30 CCGs/Primary Care divisions
- 12 ambulance NHS trusts
- 6 hospitals
- 5 welsh local health boards

From those, 89 NHS services replied and 52 agreed to disseminate our survey within the study time frame via trust intranet, email, twitter and weekly news bulletin (as shown below). We also disseminated our survey via social media: text messages, twitter, facebook advertising, as well as in a website we created for our study ([www.covidastudy.com](http://www.covidastudy.com)). The study was also reported in a Radio 4 news programme which may have further encouraged participation.

**NHS HEALTHCARE services that disseminated the survey amongst their staff**

Barnet, Enfield and Haringey Mental Health NHS Trust.

Camden & Islington NHS Foundation trust

West Hertfordshire Hospitals NHS Trust & Watford General Hospital

North Middlesex University Hospital NHS Trust

Royal National Orthopaedic Hospital NHS Trust

North East London NHS Foundation Trust

Central and North West London NHS Foundation Trust

Whittington Health NHS Trust

Bolton NHS Foundation Trust

United Lincolnshire Hospitals NHS Trust

Barts Health NHS Trust

The Princess Alexandra Hospital NHS Trust

Harrogate & District NHS Foundation Trust

Frimley Health NHS Foundation Trust

Bradford District and Craven Clinical Commissioning Group (CC)

Medway NHS Foundation Trust NSH

LNW Primary Care NIHR

Bradford Teaching Hospitals NHS Foundation Trust

Cambridgeshire Community Services NHS Trust

Cambridgeshire and Peterborough NHS Foundation Trust

Sussex Community NHS Foundation Trust

Norfolk and Suffolk Primary and Community Care Research Office /NHS Clinical Commissioning Group.Primary care and NHS trust

Airedale NHS Foundation Trust

Lewisham & Greenwich NHS Trust

Surrey and Borders Partnership NHS Foundation Trust

Mid and South Essex University Hospitals Group

University Hospitals of North Midlands NHS Trust

Queen Victoria Hospital NHS Foundation Trust

Birmingham and Solihull Mental Health NHS Trust

Hertfordshire Community NHS Trust & Hertfordshire Partnership University NHS Foundation Trust

Powys Teaching Health Board

Liverpool Women’s Hospital

Norfolk and Norwich University Hospitals NHS Foundation Trust

Gloucestershire Health and Care NHS Foundation Trust

Hounslow and Richmond Community Healthcare NHS Trust

Royal Berkshire NHS Foundation Trust

The Dudley Group NHS Foundation Trust

Tees, Esk and Wear Valleys NHS Foundation Trust,

The Rotherham NHS Foundation Trust

Northern Care Alliance NHS Group Comprising the Care Organisations of Salford, Bury & Rochdale, Oldham and North Manchester

Worcestershire Acute Hospitals NHS Trust

NHS Orkney

Gloucester hospital NHS foundation trust

Dorset Healthcare University NHS Foundation Trust

Cambridge University Hospitals NHS Foundation Trust

Royal Surrey County Hospital NHS Foundation Trust

Cwm Taf Morgannwg University Health Board| Royal Glamorgan Hospital

Tameside and Glossop Integrated Care NHS Foundation Trust

East Cheshire NHS Trust

Kingston Hospital NHS Foundation Trust

Shrewsbury and Telford hospital NHS trust

Kent and Medway NHS and Social Care Partnership Trust

**S3 Covida Study Survey**

The survey is divided in to five sections: (A) Demographics and Role; (B) Workplace (C) Risk Management; (D) Experience of traumatic and stressful events; (E) Protective factors. Subsidiary or follow-up questions (shaded) were not analysed as potential predictor variables. In addition, section F shows pre-COVID and during-COVID questions about wellbeing, worries and views about work.

A. **Demographics and Role**

|  | **Question** | **Coding** |
| --- | --- | --- |
| 1 | Gender? | 1. Male 2. Female |
| 2 | Your age? | 1. Under 25 2. 25-34 3. 35-44 4. 45-54 5. 55-64 6. 65 and above |
| 3 | What is your ethnicity?  Choice of 1. English/Welsh/Scottish/Northern Irish/ British 2. Irish 3. Gypsy or Irish Traveller 4. Any other white background 5. Mixed – White and Black Caribbean 6. Indian 7. Pakistani 8. Bangladeshi 9. Chinese 10. Any other Asian background 11. African 12. Caribbean 13. Any other Black/ African/ Caribbean background 14. Arab 15. Any other ethnic group 16. I prefer not to say | Ethnicity was coded as BAME (not white) vs white. |
| 4 | What is your relationship status? Choice of 1. Single 2. In a relationship (not living together) 3. In a relationship (living together) 4. Married 5. Divorced 6. Separated 7. Separated 8. Prefer not to say 9. Other (please specify, or skip) | Those in a relationship were combined (2/3/4). 5 and 6 were combined. |
| 5 | Are you responsible for the care of children? Choice of | Yes No |
|  | What setting do you USUALLY work in? Choice of 1. NHS Hospital 2. NHS GP surgery 3. NHS Other community team 4. NHS Mental Health Trust 5. NHS Ambulance service 6. A private hospital e.g. Priory 7. Care home 8. Nursing home 9. Other health care setting (please specify) 10 I do NOT work in health care. |  |
|  | Is your workplace managed by: Choice of 1. NHS 2. A charity 3. A private company |  |
|  | Is your job permanent, fixed term, or are you in a locum post or training programme? Choice of 1. Fixed term 2. Permanent 3. Locum 4. In a training programme 5. None of the above |  |
| 6 | What is your USUAL role? (if you have re-deployed, there is a separate question about that later) Choice of 1. Nurse 2. Midwife 3. Ambulance service personnel (e.g. paramedic. Please specify: ) 4. Mental Health Nurse 5. Health Assistant 6. Allied Healthcare Professional (e.g. OT…). Please specify: 7. Carer in a care home setting 8. A final year medical student with an early provisional registration 9. FY Doctor 10 CT Doctor 11. GP 12. Senior Doctor/ Speciality Doctor 13. Consultant 14. Management – if so what role 15. I had left the NHS but have returned to help with COVID outbreak 16. Other health worker role (please specify) 17. Administration 18. Other NON-health worker role | Groups were collapsed in the following way: Nurses (1/2/4), Doctors (9/10/11/12/13), Allied (6), Management (14), Ineligible (17/18), Other HCW (Ambulance and HA) (3/5/8/15/16 |
|  | What is your speciality ? ( or skip if none) |  |
| 7 | Where is your place of work?  Choice of 1. England – outside London 2. England – inside London 3. Northern Ireland 4. Wales 5. Scotland 6. Another country. | London vs Outside London |
|  | What NHS trust do you work for, or care/nursing home name? |  |
|  | Do you USUALLY (i.e. before being redeployed if you have work with in-patients or out-patients?) Choice of 1. In-patients 2. Out-patients 3. Both |  |
| 8 | Do you USUALLY (i.e. before being redeployed if you have) care for / treat the elderly (i.e. in a care home/nursing home, or care of the elderly in-patient unit)? | Yes No |
|  | How many hours are you currently working PER WEEK NOW relative to your usual PRE-COVID working week. (Move the green slider circle left/right. Negative numbers mean you're working LESS. 0 is the same. Positive numbers mean you're working MORE). | Number |
| 9 | Have you been re-deployed from your usual role and dept. to a different role OR dept. (i.e. more directly treating COVID patients) because of the COVID outbreak? | Yes No |
| 10 | I consider myself to be on the 'frontline' in combatting Covid-19. This can be defined as being directly engaged in clinical activities of diagnosing, treating, or providing nursing care to patients with elevated temperature/persistent cough or patients with confirmed COVID-19? | Yes No |
| 11 | Do you have any of the following. These are the NHS England risk factors for coronavirus. If you take medication for these please enter the medication in the text field.  Choice of 1. Are you 70 or older 2. Are pregnant 3. Long conditions, such as asthma, COPD, emphysema, or bronchitis 4. Heart disease, such as heart failure. 5. Chronic kidney disease 6. Liver disease, such as hepatitis 7. Conditions affecting the brain and nerves, such as Parkinson’s disease, motor neurone disease, multiple sclerosis (MS), a learning disability or cerebral palsy 8. Diabetes 9. Problems with your spleen – for example, sickle cell disease or if you have had your spleen removed 10. A weakened immune system as the result of conditions such as HIV and AIDS or medicines such as steroid tables or chemotherapy 11. Being very overweight (having a BMI of 40 or above) 12. None of these | Confirmation of any factor = ‘at risk’ vs ‘not at risk’. |
| 12 | Do you have any diagnosed mental health issues at this time? (A list was provided of MH diagnoses).  Is the diagnosis indicative of one of the following disorders? Choice of 1. Anxiety 2. Depression 3. OCD: obsessive compulsive disorder 4. PTSD: post traumatic stress disorder 5. Bipolar affective disorder 6. Eating disorder 7. Psychotic illness (e.g. schizophrenia) 8. Other (please specify) : 9. I do not wish to answer this | Yes No |
|  | Are you taking any of the following medications or treatments? IF so, please specify the medication you are taking.  Choice 1. Antidepressants 2. Sleeping tablets 3. Medication for anxiety – “anxiolytics” 4. Antipsychotics 5. ‘Talking therapies’ e.g. CBT (cognitive behavioural therapy) or counselling etc. 6. I take none of the above 7. I do not wish to answer this. |  |

B. **Experience of traumatic and stressful events**

| 13 | How many of your colleagues have been, or are currently, sick from COVID? | 0 vs 1+ |
| --- | --- | --- |
| 14 | How many of your colleagues have unfortunately died due to COVID? | 0 vs 1+ |
| 15 | How many patients in your healthcare setting have unfortunately died from COVID? | 0 vs 1+ |
| 16 | Have you lost someone you were close to from COVID?  Choice of 1. Yes, a close family member/s 2. Yes, a friend/s 3. Yes, family and (a) friend (s) 4. No | 0 vs Yes (any) |
|  | Are you doing each of these more, the same, or less now since COVID than usual?   a. Delivering bad news 17 b. Making end of life care plans 18 c. Signing Do Not Resuscitate orders 19 d. Verifying death 20 e. Signing a death certificate 21 f. Providing aftercare for a deceased patient 22 g. Withdrawing treatment e.g. oxygen, ventilator from a patient as it was deemed to be futile h. Cardio-pulmonary resuscitation 23 i. Patients asking you if they are going to die 24 j. Patients asking you if you can let them die/end their life 25 | Each coded as: more vs not more |
| 26 | In your position are you responsible (whether you have yet or not) for making important decisions about whether COVID patients should receive treatment? (99)  Follow-up questions.   a. Have you had to take such a decision to date? (If so, you can provide more info, but if not please skip) (100) Choice of 1. Yes – and I had to deny treatment to a patient 2. Yes – but I was able to decide to provide treatment to the patient 3. No 4. I do not want to answer this b. Did you / do you have the support of an ethical panel in your work place to make such decisions (114) Choice of 1. Yes 2. No | Yes No  As shown, left.  Yes No |
|  | Since COVID started have you experienced the following stressful or traumatic event(s) (you can choose multiple options if relevant): Choice of 1. A patient / patients dying of COVID 2. A patient / patients extremely unwell due to COVID 3. A colleague (s) became very sick from COVID 4. A colleague died from COVID 5. I had to make clinical decisions which were stressful or traumatising 6. Denying visits from friends/family 7. Other 8. None | Confirmation of any = “experienced trauma” vs “not” |
| 27 | Did you witness / know about the COVID-relevant stressful or traumatic event(s) at work, or were you a part of it (e.g. treating the patient). (you can choose multiple options if relevant) | 1. Witnessed it 2. I was part of the treatment 3. Know about it 4. Other |
| 28 | Taken together, would you say that you have thus experienced (either witnessed, been part of, or just "know of") COVID-relevant stressful or traumatic events at work since the outbreak began. Choice 1. Yes 2. No. | Filter for  IES-R |

C. **Workplace**

| 29 | Is your team currently missing personnel who are off because they are sick from COVID or in isolation due to actual or suspect COVID symptoms. If so, approximately what proportion of the team is missing? Choice of 1. None 2. 1-10% 3. 11-20% 4. 21 – 30% 5. 31 – 40% 6. 41 – 50% 7. >51% | None vs 1-20% vs 21%+ |
| --- | --- | --- |
| 30 | Has this absence caused extra workload for you? Choice of 1. No 2. Yes, some 3. Yes a lot | Yes (Yes a lot and some) No |
| 31 | Do you think your workplace took *sufficient* action to be prepared for the COVID outbreak before it started? | Yes No |
| 32 | Do you think there was in fact sufficient time to become adequately prepared from e.g. seeing things develop in China/Italy/Spain, but this was not done? | Yes No |
| 33 | Do you think the information you received about COVID is sufficient? | Yes No |
| 34 | Have you received any specific training around COVID? 1. Yes 2. Partly 3. No | Yes (Yes + partly) vs No |

D. **Management of risk**

| **35** | Do you think enough is being done now to reduce the risk of your exposure to COVID infection at work? Choice of 1. Yes 2. No | Yes No |
| --- | --- | --- |
| 36 | Have you had a past diagnosis, or suspected of having COVID? Or are you currently experiencing symptoms consistent with COVID? (A high temperature and/or a persistent cough) Choice of 1. In the past only - a test was positive. And now I'm fine. 2. In the past only - a test was negative. And now I'm fine 3. In the past only - suspected, but I was not tested. And now I'm fine 4. I have current symptoms - suspected COVID - and have been tested (awaiting results) 5. I have current symptoms - suspected COVID - and have NOT been tested 6. I have no current or prior symptoms suggestive of COVID 7. Other | Coded as shown left |
|  | To confirm, have you been tested for COVID at work? (i.e. with a medical testing kit). Choice of 1. Yes 2. No | Yes No |
| 37 | As you have experienced or are experiencing symptoms, have you had (or are you currently having) days off work to self-isolate due to the presence of these symptoms? Choice of 1. Yes 2. No | Yes No |
|  | How many days did you have, or are currently planned to, have off to self-isolate? (Sliding Question)  Number of days of work – slide for the correct number | Number of days |
| 38 | Have you moved out of your usual accommodation (with partner/family/friends) because of your elevated risk of passing on COVID due to being a healthcare worker? Choice of 1. Yes 2. No. | Yes No |
| 39 | Do you feel there would be / would have been lower risk of contracting COVID if there had been better planning in your workplace?  Choice of 1. Yes 2. Maybe 3. No 4. Not sure | Yes No (No, Maybe, Not sure) |

(PPE)

| 40 | Do you currently (i.e. today) have full and adequate access to the following COVID-relevant PPE safety equipment while at work, if you needed them? Choice of 1. Yes 2. Only partly 3. No. | Yes No (No and only partly) |
| --- | --- | --- |
| 41 | On general PPE availability throughout the COVID outbreak to date: before today do you generally feel that you HAVE HAD adequate PPE (Personal Protective Equipment) during the COVID outbreak? Choice of 1. No 2. Occasionally 3. Most of the time 4. All of the time 5. Not relevant to my role. | Coded as shown left |
| 42 | Have you been pressured by your manager to work without PPE despite policy being to wear it?  Choice 1. Yes 2. No | Yes No |
| 43 | Have you been asked to re-use PPE?  Choice of 1. Yes 2. No | Yes No |
| 44 | Have you refused to work without PPE?  Choice of 1. Yes 2. No | Yes No |
|  | Have you been threatened with disciplinary action / being fired for speaking out about lack of PPE or other safety measures which should protect you from COVID?  Choice of 1. Yes 2. No | Yes No |
| 45 | Changes in access to PPE:  Is *your* access to sufficient PPE getting better or worse  Choice of 1. Much worse 2. A little worse 3. Staying the same 4. A little better 5. A lot better | Coded as shown left |

E. **Protective factors**

| 46 | CD-RISC resilience scale | Total score |
| --- | --- | --- |
| 47 | Do you feel able to share the stress/anxiety COVID is causing you within your workplace? Choice of 1. Yes 2. Partly 3. No | Yes No (No and partly) |

F. **Pre COVID and during COVID. Wellbeing, worries and views about work:**

Please tell us how you *generally* felt in the 4 week period BEFORE COVID outbreak affected your workplace, and in general SINCE COVID began affecting your workplace: "not at all", “a little”, “moderately”, “quite a lot”, "very".  Did you / do you feel:

**Negative factors**

Stressed

Feeling low

Anxious

Lack Motivation

Low sleep

Hopeless

Burnout

Own Health Concerns

Family health concerns

Worry NHS under resourced

Worry management poor

Unsupported

Stress will lead to errors

Need psychological help

Want to quit

Self-harm

Suicidal

**Positive Factors**

Resilient

Feeling driven

Team is effective

Working with other teams effective

Tech confident

Remain Positive

Supported

Valued

Proud

G. **Worries – source of worry**

Taking everything together what causes you most worry and anxiety when you think about your work during this COVID outbreak? [rated from 0-10]

1. Your own mental health 2. The mental health of family and loved ones 3. Ability to support others 4. ‘Burning out’ 5. Lack of PPE 6. The *uncertainty* about whether you have COVID 7. Contracting COVID from work 7. Contracting COVID from work 8. You infecting family or friends 9. Family or friends becoming ill or dying from COVID 10. The severity of patient’s symptoms 11. The high risk of patients going to ITU 12. The high risk of patients dying 13. That things are only going to get worse 14. Lack of sleep 15. Team members getting sick or dying 16. Poor management of the crisis by your workplace 17. Poor management of the crisis by the Government

**S4: Table 1b showing additional demographics, health status and medications for the whole cohort, and stratified X^2^ (p) statistics for frontline (FL) vs non-frontline (NFL) and London vs outside London.**

|  | **Total N (%)** | **FL N (%)** | **NFL N (%)** | **p** | **London N (%)** | **Outside N (%)** | **p** |
| --- | --- | --- | --- | --- | --- | --- | --- |
| **Have children** | 1140 (41.11%) | 526 (42.97%) | 482 (39.35%) | 0.1878 | 242 (33.85%) | 897 (43.80%) | <.0001 |
| **Pregnancies** | 40 (1.44%) | 5 (0.41%) | 30 (2.45%) | <.0001 | 6 (0.84%) | 34 (1.66%) | 0.1136 |
| **Any physical comorbidities** | 2095 (75.5%) | 234 (19.1%) | 346 (28.2%) | <.0001 | 155 (26.8%) | 423 (23.5%) | 0.742 |
| lung | 378 (13.63%) | 149 (12.17%) | 175 (14.29%) | 0.1229 | 111 (15.52%) | 266 (12.99%) | 0.089 |
| heart | 49 (1.77%) | 14 (1.14%) | 26 (2.12%) | 0.0561 | 13 (1.82%) | 36 (1.76%) | 0.9161 |
| kidney | 17 (0.61%) | 5 (0.41%) | 9 (0.73%) | 0.2844 | 3 (0.42%) | 13 (0.63%) | 0.5138 |
| liver | 16 (0.58%) | 7 (0.57%) | 9 (0.73%) | 0.6171 | 5 (0.70%) | 11 (0.54%) | 0.6227 |
| brain | 30 (1.08%) | 7 (0.57%) | 17 (1.39%) | 0.0404 | 10 (1.40%) | 20 (0.98%) | 0.3485 |
| Diabetes | 84 (3.03%) | 30 (2.45%) | 41 (3.35%) | 0.1864 | 23 (3.22%) | 61 (2.98%) | 0.7494 |
| spleen | 8 (0.29%) | 3 (0.25%) | 4 (0.33%) | 0.7059 | 1 (0.14%) | 7 (0.34%) | 0.3869 |
| Immunosuppress such as AIDS or cancer | 65 (2.34%) | 12 (0.98%) | 42 (3.43%) | <.0001 | 21 (2.94%) | 44 (2.15%) | 0.231 |
| Overweight | 120 (4.33%) | 52 (4.25%) | 53 (4.33%) | 0.9239 | 19 (2.66%) | 100 (4.88%) | 0.0116 |
| **Mental health diagnosis** | 473 (17.06%) | 194 (15.85%) | 213 (17.39%) | 0.4378 | 101 (14.13%) | 372 (18.16%) | 0.0681 |
| Anxiety | 324 (11.68%) | 132 (10.78%) | 141 (11.51%) | 0.5682 | 65 (9.09%) | 259 (12.65%) | 0.011 |
| Depression | 307 (11.07%) | 129 (10.54%) | 138 (11.27%) | 0.5643 | 69 (9.65%) | 238 (11.62%) | 0.1488 |
| OCD | 20 (0.72%) | 7 (0.57%) | 11 (0.90%) | 0.3449 | 5 (0.70%) | 15 (0.73%) | 0.9283 |
| PTSD | 52 (1.88%) | 26 (2.12%) | 19 (1.55%) | 0.291 | 6 (0.84%) | 46 (2.25%) | 0.0171 |
| Bipolar | 13 (0.47%) | 5 (0.41%) | 6 (0.49%) | 0.7635 | 6 (0.84%) | 7 (0.34%) | 0.0943 |
| Eating | 15 (0.54%) | 8 (0.65%) | 4 (0.33%) | 0.2465 | 2 (0.28%) | 13 (0.63%) | 0.266 |
| Psychotic | 1 (0.04%) | 1 (0.08%) | 0 (0.00%) | 0.317 | 0 (0.00%) | 1 (0.05%) | 0.5545 |
| **MH meds** |  |  |  | 0.322 |  |  | 0.036 |
| Antidepressants | 379 (13.67%) | 154 (12.58%) | 173 (14.12%) |  | 85 (11.89%) | 294 (14.36%) |  |
| Sleeping tablets | 63 (2.27%) | 30 (2.45%) | 26 (2.12%) |  | 17 (2.38%) | 46 (2.25%) |  |
| Antipsychotics | 3 (0.11%) | 0 (0.00%) | 3 (0.24%) |  | 3 (0.42%) | 0 (0.00%) |  |
| Talking therapies | 0 (0%) | 0 (0%) | 0 (0%) |  | 0 (0%) | 0 (0%) |  |
| None of these | 1887 (77.1%) | 954 (77.94%) | 933 (76.16%) |  | 552 (77.2%) | 1563 (76.32%) |  |
| Not reported | 18 (0.65%) | 77 (6.29) | 82 (6.69%) |  | 57 (7.97%) | 128 (6.25%) |  |
| **Started meds due to COVID-19** | 85 (3.07%) | 40 (3.27%) | 33 (2.69%) | 0.3297 | 22 (3.08%) | 63 (3.08%) | 0.9029 |
| **Type of contract** |  |  |  | 0.032 |  |  | <.0001 |
| Fixed term | 280 (10.10%) | 128 (10.46%) | 122 (9.96%) |  | 76 (10.63%) | 204 (9.96%) |  |
| Permanent | 2298 (82.87%) | 993 (81.13%) | 1034 (84.41%) |  | 557 (77.9%) | 1736 (84.77%) |  |
| Locum | 36 (1.30%) | 16 (1.31%) | 18 (1.47%) |  | 18 (2.52%) | 17 (0.83%) |  |
| In a training programme | 98 (3.53%) | 55 (4.49%) | 31 (2.53%) |  | 58 (8.11%) | 40 (1.95%) |  |
| Not reported | 61 (2.20%) | 32 (2.61%) | 20 (1.63%) |  | 6 (0.84%) | 51 (2.49%) |  |
| **Redeployed** | 734 (26.47%) | 459 (37.50%) | 189 (29.17%) | <.0001 | 214 (29.93%) | 519 (25.34%) | 0.0469 |
| **Inpatients /outpatients** |  |  |  | <.0001 |  |  | 0.0021 |
| Inpatients | 792 (28.56%) | 534 (43.63%) | 189 (14.53%) |  | 172 (24.06%) | 619 (30.22%) |  |
| Outpatients | 1077 (38.84%) | 323 (26.39%) | 634 (51.76%) |  | 279 (39.02%) | 796 (38.87%) |  |
| Both | 773 (27.88%) | 338 (27.61%) | 332 (27.10%) |  | 233 (32.59%) | 538 (26.27%) |  |
| **Working with elderly** | 1106 (39.88%) | 679 (55.47%) | 298 (24.33%) | <.0001 | 230 (32.17%) | 875 (42.72%) | <.0001 |
| **Extra Working hours mean (sd)*** | 10.25 (16.13) | 12.63 (15.8) | 7.67 (16.3) | <.0001 | 10.51 (15.9) | 10.16 (16.2) | 0.669 |

**S5: table 7 showing frequency (N (%)) distributions and X2 statistics (p) for the additional significant predictors retained in the symptom models (shown in table 3) for the total cohort and stratified X^2^ (p) statistics for frontline (FL) vs non-frontline (NFL) and inside London vs outside London**

|  |  | **Total N** | **FL N (%)** | **NFL N (%)** | **p** | **London N (%)** | **Outside London N (%)** | **p** |
| --- | --- | --- | --- | --- | --- | --- | --- | --- |
| **Traumatic and stressful events** | **Friend(s) or family died** |  |  |  | 0.0045 |  |  | 0.134 |
|  | No | 2467 (91%) | 1075 (89.51%) | 1109 (92.8%) |  | 630 (89.62%) | 1829 (91.5%) |  |
|  | Yes | 244 (9%) | 126 (10.49%) | 86 (7.2%) |  | 73 (10.38%) | 170 (8.5%) |  |
|  | **Patient asking if dying** |  |  |  | <.0001 |  |  | 0.6607 |
|  | not more | 2188 (78.9%) | 757 (61.85%) | 1167 (95.27%) |  | 568 (79.44%) | 1611 (78.66%) |  |
|  | more | 585 (21.1%) | 467 (38.15%) | 58 (4.73%) |  | 147 (20.56%) | 437 (21.34%) |  |
|  | **Colleagues with COVID-19** |  |  |  | <.0001 |  |  | 0.001 |
|  | None | 540 (19.5%) | 132 (10.8%) | 342 (27.9%) |  | 107 (19.9%) | 431 (21%) |  |
|  | 1+ | 2233 (80.5%) | 1092 (89.2%) | 882 (72.1%) |  | 608 (85%) | 1617 (79%) |  |
|  | **Colleagues died from COVID-19** |  |  |  | <.0001 |  |  | <.0001 |
|  | None | 2378 (85.7%) | 1005 (82.1%) | 1092 (89.1%) |  | 546 (76.4%) | 1822 (89%) |  |
|  | 1+ | 395 (14.2%) | 219 (17.9%) | 133 (10.9%) |  | 169 (23.6%) | 226 (11%) |  |
|  | **Resuscitation** |  |  |  | <.0001 |  |  | 0.0107 |
|  | not more | 2639 (95.17%) | 1100 (89.87%) | 1222 (99.76%) |  | 668 (93.43%) | 1962 (95.8%) |  |
|  | more | 134 (4.83%) | 124 (10.13%) | 3 (0.24%) |  | 47 (6.57%) | 86 (4.2%) |  |
|  | **Giving bad news** |  |  |  | <.0001 |  |  | 0.3145 |
|  | not more | 2352 (84.82%) | 883 (36.06%) | 1176 (48.02%) |  | 598 (83.64%) | 1745 (85.21%) |  |
|  | more | 421 (15.18%) | 341 (13.92%) | 49 (2%) |  | 117 (16.36%) | 303 (14.79%) |  |
|  | **Aftercare deceased** |  |  |  | <.0001 |  |  | 0.0344 |
|  | not more | 2491 (89.83%) | 975 (79.66%) | 1208 (98.61%) |  | 657 (91.89%) | 1825 (89.11%) |  |
|  | more | 282 (10.17%) | 249 (20.34%) | 17 (1.39%) |  | 58 (8.11%) | 223 (10.89%) |  |
| **Workplace** | **Extra workload due to team absence** |  |  |  | <.0001 |  |  | 0.246 |
|  | No | 1317 (47.49%) | 471 (38.5%) | 707 (57.7%) |  | 323 (45.2%) | 991 (48.4%) |  |
|  | Some | 1134 (40.89%) | 597 (48.8%) | 394 (32.2%) |  | 309 (43.2%) | 820 (40.0%) |  |
|  | Yes | 254 (9.16%) | 139 (11.4%) | 84 (6.9%) |  | 70 (9.8%) | 184 (9.0%) |  |
|  | NR | 68 (2.45%) | 17 (1.4%) | 40 (3.3%) |  | 13 (1.8%) | 53 (2.6%) |  |
|  | **Is information you receive about COVID-19 sufficient?** |  |  |  | <.0001 |  |  | 0.335 |
|  | No | 559 (20.2%) | 300 (24.5%) | 186 (15.2%) |  | 133 (18.6%) | 426 (76.2%) |  |
|  | Yes | 2201 (79.3%) | 918 (75.0%) | 1035 (84.5%) |  | 580 (91.1%) | 1612 (78.7%) |  |
|  | NR | 13 (0.5%) | 6 (0.5%) | 4 (0.3%) |  | 2 (0.3%) | 10 (0.5%) |  |
|  | **Received specific training around COVID-19** |  |  |  | <.0001 |  |  | 0.989 |
|  | No | 1223 (44.1%) | 448 (36.6%) | 648 (52.9%) |  | 315 (44.1%) | 904 (44.1%) |  |
|  | Partly | 566 (20.41%) | 269 (22.0%) | 224 (18.3%) |  | 143 (20.0%) | 420 (20.5%) |  |
|  | Yes | 964 (34.76%) | 503 (41.1% | 340 (27.8%) |  | 252 (35.2%) | 710 (34.7%) |  |
|  | NR | 20 (0.72%) | 4 (0.3%) | 13 (1.1%) |  | 5 (0.7%) | 14 (0.7%) |  |
|  | **Team off sick** |  |  |  | <.0001 |  |  | 0.225 |
|  | 0% | 553 (19.9%) | 142 (11.6%) | 357 (29.1%) |  | 133 (18.6%) | 419 (20.5%) |  |
|  | 1-20% | 1813 (65.38%) | 844 (69.0% | 742 (60.6%) |  | 461 (64.5%) | 1344 (65.6%) |  |
|  | 21+% | 407 (14.68%) | 234 (19.1%) | 118 (9.6%) |  | 117 (16.4%) | 274 (13.4%) |  |
|  | NR | 16 (0.57%) | 4 (0.3%) | 8 (0.7%) |  | 4 (0.6%) | 11 (0.5%) |  |
| **Management of Risk** | **Enough being done now to reduce risk at work** |  |  |  | <.0001 |  |  | 0.463 |
|  | No | 974 (35.12%) | 518 (42.3%) | 334 (27.3%) |  | 264 (36.9%) | 705 (34.4%) |  |
|  | Yes | 1783 (64.3%) | 701 (57.3%) | 883 (72.1%) |  | 447 (62.5%) | 1333 (65.1%) |  |
|  | NR | 16 (0.6%) | 5 (0.4%) | 8 (0.7%) |  | 4 (0.6%) | 10 (0.5%) |  |
|  | **Pressure to work without PPE** |  |  |  | <.0001 |  |  | 0.7241 |
|  | No | 2230 (80.42%) | 953 (77.86%) | 1025 (83.67%) |  | 568 (79.44%) | 1655 (80.81%) |  |
|  | Yes | 362 (13.05%) | 214 (17.48%) | 92 (7.51%) |  | 98 (13.71%) | 264 (12.89%) |  |
|  | NR | 181 (6.53%) | 57 (4.66%) | 108 (8.82%) |  | 49 (6.85%) | 129 (6.3%) |  |
|  | **Pressure to re-use PPE** |  |  |  | <.0001 |  |  | 0.0001 |
|  | No | 1917 (69.13%) | 712 (58.17%) | 986 (80.49%) |  | 535 (74.83%) | 1377 (67.24%) |  |
|  | Yes | 713 (25.71%) | 482 (39.38%) | 140 (11.43%) |  | 141 (19.72%) | 570 (27.83%) |  |
|  | NR | 143 (5.16%) | 30 (2.45%) | 99 (8.08%) |  | 39 (5.45%) | 101 (4.93%) |  |
|  | **Less risk if workplace had better preparation** |  |  |  | <.0001 |  |  | 0.013 |
|  | No | 1142 (41.2%) | 425 (34.7%) | 590 (48.2%) |  | 276 (38.6%) | 864 (42.2%) |  |
|  | Maybe | 910 (32.8%) | 426 (34.8%) | 380 (31%) |  | 222 (31%) | 684 (33.4%) |  |
|  | Yes | 716 (25.8%) | 372 (30.4%) | 253 (20.7%) |  | 217 (30.3%) | 498 (24.3%) |  |
|  | NR | 5 (0.2%) | 1 (0.1%) | 1 (0.1%) |  | 0 (0%) | 2 (0.1%) |  |
| **Protective** | **Share stress** |  |  |  | 0.0261 |  |  | 0.0453 |
|  | No | 443 (15.98%) | 220 (17.97%) | 173 (14.12%) |  | 108 (15.1%) | 334 (16.31%) |  |
|  | Partly | 942 (33.97%) | 389 (31.78%) | 437 (35.67%) |  | 274 (38.32%) | 666 (32.52%) |  |
|  | Yes | 1319 (47.57%) | 581 (47.47%) | 588 (48%) |  | 318 (44.48%) | 997 (48.68%) |  |
|  | NR | 69 (2.49%) | 34 (2.78%) | 27 (2.2%) |  | 15 (2.1%) | 51 (2.49%) |  |

*PPE=Personal Protective Equipment*

**S6: Table 8 showing PPE and risk management factors for the whole cohort, and stratified X^2^ (p) statistics for frontline (FL) vs non-frontline (NFL) and London vs outside London.**

|  | **Total N** | **Total %** | **FL N** | **FL %** | **NFL N** | **NFL %** | **p** | **London N** | **London %** | **Outside London N** | **Outside London %** | **p** |
| --- | --- | --- | --- | --- | --- | --- | --- | --- | --- | --- | --- | --- |
| **Current access to PPE** |  |  |  |  |  |  | <.0001 |  |  |  |  | 0.0055 |
| No | 188 | 6.78 | 64 | 5.23 | 110 | 8.98 |  | 52 | 7.27 | 136 | 6.64 |  |
| Partly | 597 | 21.53 | 304 | 24.84 | 203 | 16.57 |  | 185 | 25.87 | 409 | 19.97 |  |
| Yes | 1871 | 67.47 | 829 | 67.73 | 831 | 67.84 |  | 447 | 62.52 | 1420 | 69.34 |  |
| NR | 117 | 4.22 | 27 | 2.21 | 81 | 6.61 |  | 31 | 4.34 | 83 | 4.05 |  |
| **Beginning of outbreak access to PPE** |  |  |  |  |  |  | <.0001 |  |  |  |  | 0.0017 |
| No | 329 | 11.86 | 168 | 13.73 | 123 | 10.04 |  | 92 | 12.87 | 236 | 11.52 |  |
| Occasionally | 297 | 10.71 | 144 | 11.76 | 104 | 8.49 |  | 86 | 12.03 | 209 | 10.21 |  |
| Most of the time | 1032 | 37.22 | 513 | 41.91 | 401 | 32.73 |  | 260 | 36.36 | 771 | 37.65 |  |
| All of the time | 670 | 24.16 | 358 | 29.25 | 229 | 18.69 |  | 140 | 19.58 | 530 | 25.88 |  |
| Not relevant | 343 | 12.37 | 9 | 0.74 | 311 | 25.39 |  | 111 | 15.52 | 230 | 11.23 |  |
| NR | 102 | 3.68 | 32 | 2.61 | 57 | 4.65 |  | 26 | 3.64 | 72 | 3.52 |  |
| **Refuse to work with no PPE** |  |  |  |  |  |  | <.0001 |  |  |  |  | 0.9083 |
| No | 2232 | 80.49 | 963 | 78.68 | 1004 | 81.96 |  | 580 | 81.12 | 1646 | 80.37 |  |
| Yes | 343 | 12.37 | 217 | 17.73 | 87 | 7.1 |  | 86 | 12.03 | 255 | 12.45 |  |
| NR | 198 | 7.14 | 44 | 3.59 | 134 | 10.94 |  | 49 | 6.85 | 147 | 7.18 |  |
| **Disciplinary action** |  |  |  |  |  |  | <.0001 |  |  |  |  | 0.6519 |
| No | 2543 | 91.71 | 1149 | 93.87 | 1101 | 89.88 |  | 651 | 91.05 | 1885 | 92.04 |  |
| Yes | 79 | 2.85 | 44 | 3.59 | 22 | 1.8 |  | 21 | 2.94 | 58 | 2.83 |  |
| NR | 151 | 5.45 | 31 | 2.53 | 102 | 8.33 |  | 43 | 6.01 | 105 | 5.13 |  |
| **Access to PPE over time** |  |  |  |  |  |  | <.0001 |  |  |  |  | 0.0075 |
| Much worse | 52 | 1.88 | 38 | 3.1 | 10 | 0.82 |  | 13 | 1.82 | 39 | 1.9 |  |
| A little worse | 245 | 8.84 | 155 | 12.66 | 68 | 5.55 |  | 68 | 9.51 | 175 | 8.54 |  |
| Staying the same | 1435 | 51.75 | 636 | 51.96 | 615 | 50.2 |  | 328 | 45.87 | 1104 | 53.91 |  |
| A little better | 526 | 18.97 | 236 | 19.28 | 231 | 18.86 |  | 160 | 22.38 | 365 | 17.82 |  |
| A lot better | 290 | 10.46 | 124 | 10.13 | 129 | 10.53 |  | 78 | 10.91 | 211 | 10.3 |  |
| NR | 225 | 8.11 | 35 | 2.86 | 172 | 14.04 |  | 68 | 9.51 | 154 | 7.52 |  |
| **Workplace took sufficient action to prepare** |  |  |  |  |  |  | 0.358 |  |  |  |  | <.0001 |
| No | 1281 | 42.92 | 536 | 42.8 | 529 | 43.2 |  | 382 | 53.4 | 833 | 40.7 |  |
| Yes | 1542 | 55.61 | 686 | 56 | 690 | 56.3 |  | 331 | 46.3 | 1206 | 58.9 |  |
| Not reported | 13 | 0.47 | 2 | 0.2 | 6 | 0.5 |  | 2 | 0.3 | 9 | 0.4 |  |
| **Had been sufficient time, but not done** |  |  |  |  |  |  | 0.292 |  |  |  |  | 0.398 |
| No | 278 | 22.92 | 114 | 21.3 | 134 | 25.3 |  | 88 | 23 | 190 | 22.8 |  |
| Yes | 935 | 77.08 | 420 | 78.4 | 393 | 74.3 |  | 294 | 77 | 639 | 76.7 |  |
| Not reported | 5 | 0.40 | 2 | 0.4 | 2 | 0.4 |  | 0 | 0 | 4 | 0.5 |  |
| **Days off work to self-isolate** |  |  |  |  |  |  | <.0001 |  |  |  |  | 0.3103 |
| No | 248 | 8.94 | 115 | 9.4 | 97 | 7.92 |  | 70 | 9.79 | 177 | 8.64 |  |
| Yes | 546 | 19.69 | 297 | 24.26 | 194 | 15.84 |  | 151 | 21.12 | 395 | 19.29 |  |
| NR | 1979 | 71.37 | 812 | 66.34 | 934 | 76.24 |  | 494 | 69.09 | 1476 | 72.07 |  |
| **Move out home due to COVID-19 outbreak** |  |  |  |  |  |  | <.0001 |  |  |  |  | 0.7459 |
| No | 2618 | 94.41 | 1126 | 91.99 | 1186 | 96.82 |  | 674 | 94.27 | 1936 | 94.53 |  |
| Yes | 125 | 4.51 | 84 | 6.86 | 26 | 2.12 |  | 35 | 4.9 | 90 | 4.39 |  |
| NR | 30 | 1.08 | 14 | 1.14 | 13 | 1.06 |  | 6 | 0.84 | 22 | 1.07 |  |

**S7: Table 9 showing traumatic and stressful events or duties for the whole cohort, and stratified X^2^ (p) statistics for frontline (FL) vs non-frontline (NFL) and London vs outside London**

|  | **Total N** | **Total %** | **FL N** | **FL %** | **NFL N** | **NFL %** | **p** | **London** | **London %** | **Outside London N** | **Outside London %** | **p** |
| --- | --- | --- | --- | --- | --- | --- | --- | --- | --- | --- | --- | --- |
| ***Traumatic events*** |  |  |  |  |  |  |  |  |  |  |  |  |
| **Patients died** |  |  |  |  |  |  | <.0001 |  |  |  |  | 0.35 |
| 0 (0) | 1310 | 47.2 | 337 | 27.5 | 810 | 66.1 |  | 327 | 45,7 | 978 | 47.8 |  |
| 1+ (1) | 1463 | 52.8 | 887 | 72.5 | 415 | 33.9 |  | 388 | 54.3 | 1070 | 52.2 |  |
| **Saw very unwell patients** |  |  |  |  |  |  | <.0001 |  |  |  |  | 0.85 |
| No (0) | 1626 | 58.6 | 350 | 28.6 | 1048 | 85.7 |  | 416 | 58.3 | 1200 | 58.7 |  |
| Yes (1) | 1149 | 41.4 | 873 | 71.4 | 175 | 14.3 |  | 298 | 41.7 | 846 | 41.3 |  |
| **Made a traumatic clinical decision** |  |  |  |  |  |  | <.0001 |  |  |  |  | 0.013 |
| No (0) | 2394 | 86.3 | 953 | 77.9 | 1146 | 93.7 |  | 596 | 83.5 | 1784 | 87.2 |  |
| Yes (1) | 381 | 13.7 | 270 | 22.10 | 77 | 6.3 |  | 118 | 16.5 | 262 | 12.8 |  |
| ***Stressful or traumatic duties**** |  |  |  |  |  |  |  |  |  |  |  |  |
| **End of life care** |  |  |  |  |  |  | <.0001 |  |  |  |  | 0.7497 |
| not more (0) | 2360 | 85.11 | 887 | 72.47 | 1180 | 96.33 |  | 611 | 85.45 | 1740 | 84.96 |  |
| more (1) | 413 | 14.89 | 337 | 27.53 | 45 | 3.67 |  | 104 | 14.55 | 308 | 15.04 |  |
| **Sign resuscitation** |  |  |  |  |  |  | <.0001 |  |  |  |  | 0.016 |
| not more (0) | 2569 | 92.64 | 1056 | 86.27 | 1203 | 98.2 |  | 648 | 90.63 | 1912 | 93.36 |  |
| more (1) | 204 | 7.36 | 168 | 13.73 | 22 | 1.8 |  | 67 | 9.37 | 136 | 6.64 |  |
| **Verifying death** |  |  |  |  |  |  | <.0001 |  |  |  |  | 0.0322 |
| not more (0) | 2649 | 95.53 | 1115 | 91.09 | 1216 | 99.27 |  | 673 | 94.13 | 1967 | 96.04 |  |
| more (1) | 124 | 4.47 | 109 | 8.91 | 9 | 0.73 |  | 42 | 5.87 | 81 | 3.96 |  |
| **Signing death cert** |  |  |  |  |  |  | <.0001 |  |  |  |  | <.0001 |
| not more (0) | 2674 | 96.43 | 1138 | 92.97 | 1218 | 99.43 |  | 671 | 93.85 | 1994 | 97.36 |  |
| more (1) | 99 | 3.57 | 86 | 7.03 | 7 | 0.57 |  | 44 | 6.15 | 54 | 2.64 |  |
| **Withdraw treatment** |  |  |  |  |  |  | NA |  |  |  |  | NA |
| not more (0) | 2566 | 100.00 | 1224 | 100 | 1225 | 100 |  | 715 | 100 | 2048 | 100 |  |
| more (1) | 207 | 0.00 | 0 | 0 | 0 | 0 |  | 0 | 0 | 0 | 0 |  |
| **Patient asked to let die** |  |  |  |  |  |  | <.0001 |  |  |  |  | 0.5613 |
| not more (0) | 2463 | 88.82 | 964 | 78.76 | 1202 | 98.12 |  | 639 | 89.37 | 1814 | 88.57 |  |
| more (1) | 310 | 11.18 | 260 | 21.24 | 23 | 1.88 |  | 76 | 10.63 | 234 | 11.43 |  |
| **Position to decide treatment** |  |  |  |  |  |  | <.0001 |  |  |  |  | 0.0489 |
| no (0) | 2458 | 88.64 | 998 | 81.54 | 1170 | 95.51 |  | 618 | 86.43 | 1833 | 89.5 |  |
| yes (1) | 307 | 11.07 | 224 | 18.3 | 54 | 4.41 |  | 96 | 13.43 | 209 | 10.21 |  |
| NR (14) | 8 | 0.29 | 2 | 0.16 | 1 | 0.08 |  | 1 | 0.14 | 6 | 0.29 |  |
| **Needing to deny treatment $** |  |  |  |  |  |  | 0.603 |  |  |  |  | 0.0764 |
| No (0) | 243 | 82.09 | 177 | 81.19 | 43 | 84.31 |  | 70 | 76.09 | 171 | 84.65 |  |
| Yes (1) | 53 | 17.91 | 41 | 18.81 | 8 | 15.69 |  | 22 | 23.91 | 31 | 15.35 |  |
| **Experience traumatic event #** |  |  |  |  |  |  | <.0001 |  |  |  |  | 0.4062 |
| No (0) | 764 | 29.37 | 113 | 9.54 | 586 | 52.32 |  | 186 | 28.14 | 576 | 29.84 |  |
| Yes (1) | 1837 | 70.63 | 1072 | 90.46 | 534 | 47.68 |  | 475 | 71.86 | 1354 | 70.16 |  |

******* *Figures are of those who do that role usually*

*$* *Question only given to a subset who answered yes on 'decision treatment' question.*

*# Note this question is about traumatic events per se, not specifically COVID-related events that determine IES-R administration.*

**S8: Chi-sq summary statistics table (showing X^2^ and p values) for factors and associated with each symptom outcome score. Shaded cells are p<.05. All factors significantly associated with the outcome score were entered into stepwise logistic regressions.**

|  |  | **GAD** | **p** | **IES-R** | **p** | **PHQ** | **p** | **PSS** | **p** |
| --- | --- | --- | --- | --- | --- | --- | --- | --- | --- |
| **Demographics and role** | Gender | 19.41 | <.0001 | 2.52 | 0.112 | 10.37 | 0.0013 | 26.38 | <.0001 |
|  | Age | 41.43 | <.0001 | 14.05 | 0.015 | 12.70 | 0.0263 | 52.51 | <.0001 |
|  | BAME | 0.32 | 0.573 | 14.88 | 0.0001 | 3.15 | 0.076 | 0.07 | 0.794 |
|  | Relationship status | 9.73 | 0.021 | 5.44 | 0.142 | 16.18 | 0.001 | 11.32 | 0.0101 |
|  | Care for children at home | 0.00 | 0.974 | 1.61 | 0.2048 | 6.24 | 0.0125 | 0.04 | 0.846 |
|  | Role | 33.35 | <.0001 | 28.30 | <.0001 | 31.64 | <.0001 | 28.61 | <.0001 |
|  | London | 12.34 | 0.0004 | 0.81 | 0.3671 | 4.82 | 0.0281 | 1.40 | 0.237 |
|  | Care for elderly at work | 0.10 | 0.755 | 13.72 | 0.0002 | 0.00 | 0.971 | 0.00 | 0.968 |
|  | Redeployed | 7.28 | 0.007 | 37.91 | <.0001 | 4.20 | 0.0404 | 8.47 | 0.0036 |
|  | Frontline HCW | 26.42 | <.0001 | 117.65 | <.0001 | 9.88 | 0.0017 | 13.15 | 0.0003 |
|  | COVID-19 health risk factors | 0.38 | 0.538 | 0.05 | 0.831 | 0.65 | 0.422 | 3.34 | 0.068 |
|  | Previous mental health diagnosis | 68.67 | <.0001 | 25.97 | <.0001 | 102.41 | <.0001 | 78.73 | <.0001 |
| **Traumatic & stressful events** | Colleague ill with COVID-19 | 11.47 | 0.0007 | 51.97 | <.0001 | 9.66 | 0.0019 | 9.25 | 0.0024 |
|  | Colleague/s died of COVID-19 | 2.92 | 0.087 | 52.99 | <.0001 | 0.00 | 0.981 | 1.70 | 0.192 |
|  | Patient/s died of COVID-19 | 4.83 | 0.028 | 63.67 | <.0001 | 3.36 | 0.0666 | 2.19 | 0.139 |
|  | Close friend or relative died of COVID-19 | 11.78 | 0.0006 | 42.52 | <.0001 | 10.54 | 0.0012 | 4.01 | 0.0452 |
|  | Delivering bad news | 3.55 | 0.060 | 110.61 | <.0001 | 3.09 | 0.079 | 2.04 | 0.153 |
|  | Making end of life care plan | 0.99 | 0.321 | 46.11 | <.0001 | 0.07 | 0.791 | 1.63 | 0.202 |
|  | Sign DNR forms | 1.76 | 0.184 | 4.71 | 0.03 | 5.11 | 0.0238 | 2.00 | 0.157 |
|  | Verifying patient death | 0.02 | 0.889 | 17.05 | <.0001 | 0.00 | 0.968 | 0.03 | 0.874 |
|  | Signing death certificates | 0.02 | 0.897 | 9.26 | 0.002 | 1.46 | 0.227 | 0.20 | 0.657 |
|  | Providing after care of the deceased | 14.87 | 0.0001 | 90.78 | <.0001 | 17.60 | <.0001 | 5.09 | 0.024 |
|  | Giving CPR | 12.07 | 0.0005 | 93.72 | <.0001 | 2.34 | 0.126 | 2.23 | 0.135 |
|  | Patient asking if they are dying | 21.02 | <.0001 | 168.57 | <.0001 | 13.18 | 0.0003 | 11.00 | 0.0009 |
|  | Patient asking to let them die | 19.97 | <.0001 | 134.46 | <.0001 | 11.96 | 0.0005 | 13.72 | 0.0002 |
|  | Responsible for decision-making | 3.35 | 0.067 | 6.12 | 0.0134 | 4.05 | 0.044 | 0.96 | 0.326 |
|  | Experienced trauma | 15.15 | <.0001 | 108.22 | <.0001 | 5.89 | 0.0152 | 5.69 | 0.0171 |
| **Workplace** | Team staff absence due to COVID-19 | 20.69 | <.0001 | 61.25 | <.0001 | 23.76 | <.0001 | 30.56 | <.0001 |
|  | Extra workload due to staff absence | 18.38 | <.0001 | 56.22 | <.0001 | 12.37 | 0.0004 | 19.53 | <.0001 |
|  | Insufficient workplace action in order to prepare | 27.08 | <.0001 | 27.29 | <.0001 | 15.99 | <.0001 | 22.47 | <.0001 |
|  | Insufficient COVID-19 information | 66.08 | <.0001 | 58.74 | <.0001 | 52.93 | <.0001 | 53.48 | <.0001 |
|  | Insufficient COVID-19 specific training | 13.64 | 0.0002 | 0.62 | 0.432 | 10.22 | 0.0014 | 9.42 | 0.0021 |
|  | Lower risk if better prepared | 39.52 | <.0001 | 60.87 | <.0001 | 36.21 | <.0001 | 43.29 | <.0001 |
|  | HCW COVID-19 diagnosis or symptoms | 15.16 | 0.019 | 46.22 | <.0001 | 7.16 | 0.307 | 16.47 | 0.0114 |
|  | Self-isolation | 6.36 | 0.012 | 19.62 | <.0001 | 2.50 | 0.114 | 3.94 | 0.0472 |
| **Management of Risk** | Not enough being done now to reduce risk | 72.94 | <.0001 | 66.24 | <.0001 | 46.04 | <.0001 | 39.27 | <.0001 |
|  | Moved out own home to lower family risk | 4.47 | 0.035 | 24.24 | <.0001 | 4.14 | 0.0419 | 2.37 | 0.124 |
|  | Lower current access to PPE | 19.83 | <.0001 | 29.71 | <.0001 | 11.26 | 0.0008 | 9.95 | 0.0016 |
|  | Lower general access PPE | 40.01 | <.0001 | 43.29 | <.0001 | 34.53 | <.0001 | 32.78 | <.0001 |
|  | Pressured to work with no PPE | 39.77 | <.0001 | 53.58 | <.0001 | 40.42 | <.0001 | 44.94 | <.0001 |
|  | Asked to reuse PPE | 36.16 | <.0001 | 72.43 | <.0001 | 19.02 | <.0001 | 29.74 | <.0001 |
| **Protective** | CD-RISC resilience score | 147.94 | <.0001 | 7.65 | 0.054 | 111.75 | <.0001 | 150.88 | <.0001 |
|  | Able to share stress/anxiety in workplace | 50.24 | <.0001 | 19.21 | <.0001 | 35.02 | <.0001 | 45.71 | <.0001 |

*PPE=Personal Protective Equipment.*

*CD-RISC=Connor-Davidson Resilience Scale*

*HCW=Healthcare worker*

**S9: Pre- and during-COVID-19 ratings for ‘negative’ and ‘positive’ factors (scored 1-5), and effect size of change (time). Also shown are group (Frontline/Non-frontline) x time (pre / during COVID-19) interaction effects (and description of which group showed the greatest change) from between-groups repeated-measures ANOVA.**

|  |  |  |  |  |  |  |  |  | **Group x time interaction** | | |
| --- | --- | --- | --- | --- | --- | --- | --- | --- | --- | --- | --- |
|  | **Pre COVID-19** | **sd** | **During COVID-19** | **sd** | **F** | **p** | **partial eta** | **Pre COVID-19 difference t(2447), p** | **F** | **p** | **Change magnitude** |
| **Negative factors** |  |  |  |  |  |  |  |  |  |  |  |
| Stressed | 2.29 | 0.94 | 3.30 | 1.1 | 1984 | <.0001 | 0.42 | 3.64, p<.001 (FL<NFL) | 63.2 | <.001 | FL>NFL |
| Feeling low | 1.64 | 0.89 | 2.46 | 1.2 | 1627 | <.0001 | 0.37 | 1.53, 0.13 | 37.9 | <.001 | FL>NFL |
| Anxious | 1.89 | 0.96 | 2.96 | 1.25 | 2345 | <.0001 | 0.46 | 1.43, 0.15 | 36.7 | <.001 | FL>NFL |
| Lack Motivation | 1.66 | 0.89 | 2.40 | 1.26 | 956 | <.0001 | 0.26 | 0.87, 0.39 | 1.34 | 0.247 |  |
| Low sleep | 1.92 | 1.03 | 2.86 | 1.35 | 1531 | <.0001 | 0.36 | 0.45, 0.65 | 31.0 | <.001 | FL>NFL |
| Hopeless | 1.35 | 0.75 | 2.03 | 1.24 | 1065 | <.0001 | 0.28 | 0.37, 0.71 | 44.4 | <.001 | FL>NFL |
| Burnout | 1.81 | 1.02 | 2.58 | 1.38 | 911 | <.0001 | 0.25 | 1.07, 0.29 | 81.1 | <.001 | FL>NFL |
| Own Health Concerns | 1.62 | 0.9 | 2.86 | 1.35 | 2473 | <.0001 | 0.47 | 3.07, 0.002 (FL<NFL) | 91.7 | <.001 | FL>NFL |
| Family health concerns | 1.85 | 0.93 | 3.54 | 1.2 | 5796 | <.0001 | 0.68 | 0.88, 0.38 | 46.8 | <.001 | FL>NFL |
| Worry NHS under resourced | 2.58 | 1.15 | 3.55 | 1.25 | 1898 | <.0001 | 0.41 | 3.08, 0.002 (FL<NFL) | 13.8 | <.001 | FL>NFL |
| Worry management poor | 2.27 | 1.13 | 3.11 | 1.34 | 1519 | <.0001 | 0.35 | 1.11, 0.27 | 22.4 | <.001 | FL>NFL |
| Unsupported | 1.82 | 1.04 | 2.35 | 1.38 | 616 | <.0001 | 0.18 | 1.78, 0.08 | 40.8 | <.001 | FL>NFL |
| Stress will lead to errors | 1.66 | 0.93 | 2.37 | 1.29 | 1197 | <.0001 | 0.30 | 1.65, 0.10 | 56.8 | <.001 | FL>NFL |
| Need psychological help | 1.40 | 0.81 | 1.89 | 1.15 | 753 | <.0001 | 0.21 | 2.52, 0.01 (FL<NFL) | 47.8 | <.001 | FL>NFL |
| Want to quit | 1.63 | 1.02 | 2.22 | 1.39 | 600 | <.0001 | 0.18 | 2.78, 0.005 (FL<NFL) | 39.5 | <.001 | FL>NFL |
| Self-harm | 1.08 | 0.37 | 1.16 | 0.57 | 92.9 | <.0001 | 0.03 | 0.16, 0.87 | 7.5 | 0.006 | FL>NFL |
| Suicidal | 1.07 | 0.36 | 1.15 | 0.56 | 90.5 | <.0001 | 0.03 | 0.51, 0.61 | 6.8 | 0.009 | FL>NFL |
| **Positive factors** |  |  |  |  |  |  |  |  |  |  |  |
| Resilient | 3.75 | 1 | 3.25 | 1.08 | 663 | <.0001 | 0.19 | 2.24, 0.025 (FL>NFL) | 13.5 | <.001 | FL>NFL |
| Feeling driven | 3.91 | 0.99 | 3.52 | 1.15 | 332 | <.0001 | 0.11 | 1.22, 0.22 | 1.34 | 0.25 |  |
| Team is effective | 3.80 | 1.02 | 3.71 | 1.17 | 23.0 | <.0001 | 0.01 | 3.12, 0.002 (FL>NFL) | 0.65 | 0.42 |  |
| Working with other teams effective | 3.64 | 1.05 | 3.69 | 1.1 | 5.44 | 0.02 | 0.00 | 1.87, 0.06 | 29.5 | <.001 | FL>NFL |
| Tech confident | 3.08 | 1.28 | 3.50 | 1.2 | 466 | <.0001 | 0.14 | 2.85, 0.004 (FL>NFL) | 64.1 | <.001 | NFL>FL |
| Remain Positive | 3.80 | 0.93 | 3.18 | 1.07 | 1029 | <.0001 | 0.27 | 2.55, 0.01 (FL>NFL) | 23.3 | <.001 | FL>NFL |
| Supported | 3.58 | 1.1 | 3.32 | 1.25 | 198 | <.0001 | 0.07 | 0.06, 0.95 | 10.8 | 0.001 | FL>NFL |
| Valued | 3.32 | 1.19 | 3.16 | 1.32 | 53.0 | <.0001 | 0.02 | 1.63, 0.10 | 0.11 | 0.74 |  |
| Proud | 4.06 | 1.02 | 4.16 | 1.08 | 30.0 | <.0001 | 0.01 | 0.49, 0.62 | 0.19 | 0.19 |  |

**S10. Ratings of worry across a range of factors ordered by degree of worry for the total cohort as well as a statistical comparison of worries of frontline (FL) and non-frontline (NFL) workers (t, p).**

|  | **All respondents** | ***sd*** | **NFL mean**  **(sd)** | **FL mean**  **(sd)** | **t** | **p** |
| --- | --- | --- | --- | --- | --- | --- |
| Family / friend becoming ill or dying from COVID-19 | 7.46 | *2.92* | 7.07 (0.889) | 7.81 (2.77) | 6.623 | <0.001 |
| Worry you will infect family or friends | 6.94 | *3.09* | 6.23 (0.96) | 7.56 (2.87) | 11.23 | <0.001 |
| Poor management by the Government | 6.53 | *3.17* | 6.3 (1.088) | 6.71 (3.17) | 3.22 | 0.001 |
| High risk of patients dying | 5.80 | *3.17* | 5.2 (1.079) | 6.33 (3.02) | 8.55 | <0.001 |
| Contracting COVID-19 from work | 5.71 | *3.27* | 5.06 (1.075) | 6.27 (3.18) | 9.369 | <0.001 |
| Team members getting sick or dying | 5.57 | *3.10* | 4.99 (1.042) | 6.04 (3.06) | 8.367 | <0.001 |
| Things are going to get worse | 5.38 | *3.23* | 4.86 (1.099) | 5.78 (3.22) | 6.973 | <0.001 |
| Severity of patients’ symptoms | 5.24 | *3.11* | 4.5 (1.036) | 5.84 (2.98) | 10.503 | <0.001 |
| Ability to support others | 5.17 | *2.83* | 5.01 (1.004) | 5.25 (2.85) | 2.063 | 0.039 |
| ‘Burning out' | 5.14 | *3.23* | 4.47 (1.11) | 5.55 (3.19) | 8.173 | <0.001 |
| Lack of PPE | 5.11 | *3.37* | 4.45 (1.096) | 5.53 (3.3) | 7.876 | <0.001 |
| High risk of patients going to ITU | 5.03 | *3.17* | 4.55 (1.096) | 5.41 (3.14) | 6.388 | <0.001 |
| Mental health of family and loved ones | 5.03 | *2.81* | 4.75 (1.001) | 5.3 (2.86) | 4.871 | <0.001 |
| Worry about lack of sleep | 4.76 | *3.33* | 4.08 (1.129) | 5.14 (3.28) | 7.534 | <0.001 |
| Uncertainty about having COVID-19 | 4.67 | *3.34* | 4.04 (1.14) | 5.2 (3.39) | 8.231 | <0.001 |
| Worry about own mental health | 4.22 | *2.88* | 3.79 (0.988) | 4.52 (2.89) | 6.193 | <0.001 |
| Poor management by your workplace | 4.16 | *3.33* | 3.54 (1.089) | 4.6 (3.37) | 7.492 | <0.001 |

*PPE = Personal Protective Equipment; ITU=Intensive Treatment Unit*

**Supplementary Figure 1. Figures showing change in low mood (left) and feeling anxious (centre) and feeling stressed (right), from pre to during COVID for frontline and non-frontline HCW. Error bars are 95% CI.**

| 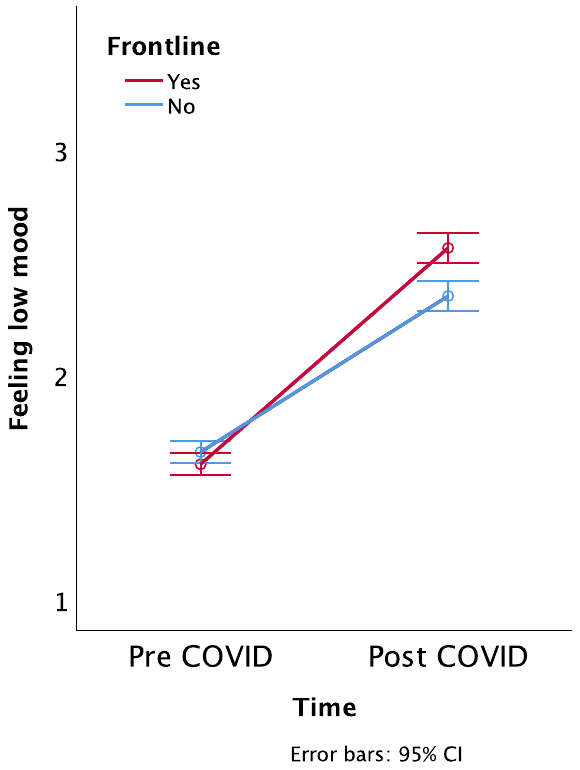 | 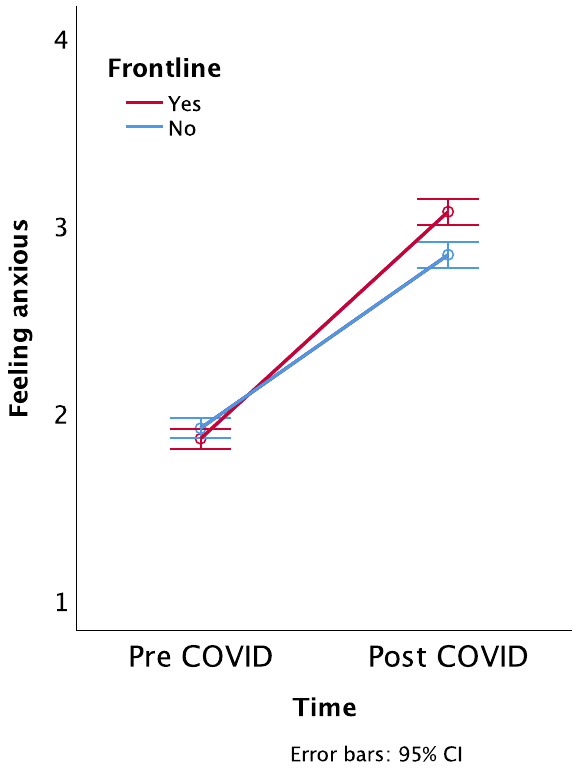 | 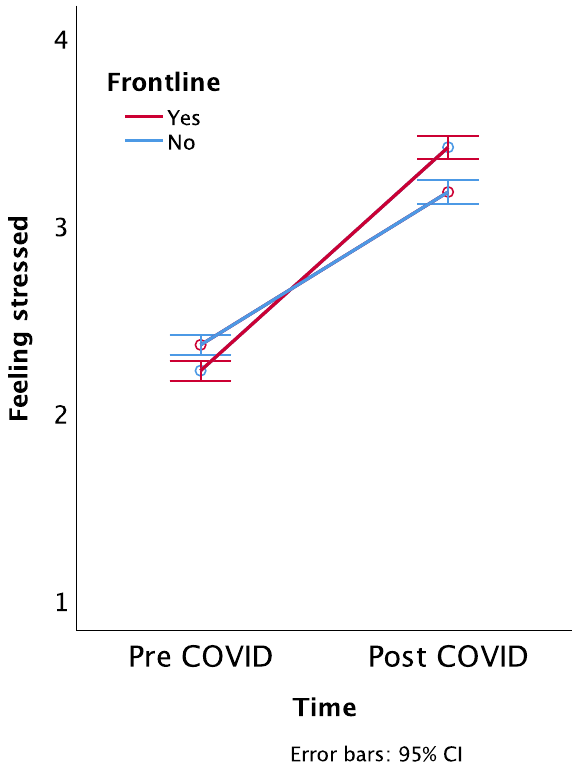 |
| --- | --- | --- |
